# Artificial Intelligence and Machine Learning in Cancer Related Pain: A Systematic Review

**DOI:** 10.1101/2023.12.06.23299610

**Authors:** Vivian Salama, Brandon Godinich, Yimin Geng, Laia Humbert-Vidan, Laura Maule, Kareem A. Wahid, Mohamed A. Naser, Renjie He, Abdallah S.R. Mohamed, Clifton D. Fuller, Amy C. Moreno

## Abstract

**Background/objective:** Pain is a challenging multifaceted symptom reported by most cancer patients, resulting in a substantial burden on both patients and healthcare systems. This systematic review aims to explore applications of artificial intelligence/machine learning (AI/ML) in predicting pain-related outcomes and supporting decision-making processes in pain management in cancer.

**Methods:** A comprehensive search of Ovid MEDLINE, EMBASE and Web of Science databases was conducted using terms including “Cancer”, “Pain”, “Pain Management”, “Analgesics”, “Opioids”, “Artificial Intelligence”, “Machine Learning”, “Deep Learning”, and “Neural Networks” published up to September 7, 2023. The screening process was performed using the Covidence screening tool. Only original studies conducted in human cohorts were included. AI/ML models, their validation and performance and adherence to TRIPOD guidelines were summarized from the final included studies.

**Results:** This systematic review included 44 studies from 2006-2023. Most studies were prospective and uni-institutional. There was an increase in the trend of AI/ML studies in cancer pain in the last 4 years. Nineteen studies used AI/ML for classifying cancer patients’ pain development after cancer therapy, with median AUC 0.80 (range 0.76-0.94). Eighteen studies focused on cancer pain research with median AUC 0.86 (range 0.50-0.99), and 7 focused on applying AI/ML for cancer pain management decisions with median AUC 0.71 (range 0.47-0.89). Multiple ML models were investigated with. median AUC across all models in all studies (0.77). Random forest models demonstrated the highest performance (median AUC 0.81), lasso models had the highest median sensitivity (1), while Support Vector Machine had the highest median specificity (0.74). Overall adherence of included studies to TRIPOD guidelines was 70.7%. Lack of external validation (14%) and clinical application (23%) of most included studies was detected. Reporting of model calibration was also missing in the majority of studies (5%).

**Conclusion:** Implementation of various novel AI/ML tools promises significant advances in the classification, risk stratification, and management decisions for cancer pain. These advanced tools will integrate big health-related data for personalized pain management in cancer patients. Further research focusing on model calibration and rigorous external clinical validation in real healthcare settings is imperative for ensuring its practical and reliable application in clinical practice.

## Introduction

According to The International Association for the Study of Pain, pain is a subjective unpleasant sensation and distressing sensory or emotional experience related to tissue damage that can vary in intensity and duration [1, 2]. In cancer, pain represents a pervasive issue affecting a large proportion of cancer patients which significantly impacts their quality of life (QoL) and increases morbidity as well as mortality rates [2, 3]. More than 55% of cancer patients, especially those with advanced stages and metastasis, complain from pain [4, 5]. Additionally, 30% of cancer patients experience chronic pain, either because of the cancer itself or due to cancer treatment such as surgery, chemotherapy (CT), and radiation therapy (RT) [6]. The burden of pain in cancer patients is substantial, impacting their physical, psychological, and social well-being [4]. Moreover, untreated or poorly managed pain can lead to decreased QoL, functional impairment, and increased opioids prescription and healthcare costs [4–7]. Pain in people with cancer can manifest in various forms, including acute or chronic. Acute pain is usually temporary but severe while chronic pain can persist and last more than 3 months after starting point of pain throughout the cancer journey in cancer survivors, [8, 9].

The management of pain in cancer patients is a complex evolving field. It involves a multimodal approach that may include pharmacological interventions, such as opioids, non-opioid analgesics and adjuvant medications, as well as non-pharmacological approaches like physical therapy, psychotherapy, and interventional techniques [10]. Pain management in cancer patients follows the widely recognized and recommended World Health Organization (WHO) analgesics ladder, which consists of a three-step approach guiding healthcare providers in selecting and prescribing appropriate pain medications based on the severity of the pain, with the aim of providing effective pain relief while minimizing side effects. [11]. Overuse of unneeded high doses of opioids raises the risks of opioids’ side effects and chronic abuse rates, which negatively affects patients’ QoL [5]. Striking the right balance between pain relief and avoiding opioid-related adverse effects and potential abuse is a significant challenge in cancer pain management [4, 5, 10].

While healthcare providers follow the WHO pain management ladder, effectively controlling pain in cancer patients remains a significant challenge. There is an unmet need for accurate guidelines to help clinicians predict, classify, and manage pain in cancer patients. Novel tools are necessary to support clinicians in risk stratification and personalized pain management, particularly in the context of cancer-related pain.

In recent years, the healthcare industry has witnessed a surge in the adoption of artificial intelligence (AI) and machine learning (ML) tools. These technologies have demonstrated their potential to transform healthcare delivery, diagnosis, and treatment decision-making [12]. AI/ML tools can analyze vast amounts of data, identify patterns, and provide predictive insights, thereby aiding healthcare professionals in making informed decisions [12, 13]. ML is a subfield of AI that includes systems that ‘learn from experience (E) with respect to some class of tasks (T) and performance measure (P), if its performance at tasks T, as measured by P, improves with experience E’ [14, 15].

Neural Networks are a subtype of ML algorithms, which can have different degrees of complexity and evolve into the so-called deep learning (DL) algorithms when they involve a large number of layers. Both ML and DL algorithms can be classed into supervised, unsupervised and reinforcement learning based on the way in which training data is presented to the model [14] (Figure 1). In supervised learning, the model is expected to learn the mapping between the training inputs and outputs presented during training to then be able to predict the outputs on an unseen input dataset. In unsupervised learning, the model is presented with unlabeled data and expected to learn the patterns and correlations by itself based on the input data only. In reinforcement learning, the algorithm learns to map inputs to actions by maximizing (or minimizing) a reward (or punishment) action evaluation signal. Additionally, AI algorithms can be grouped based on the task they are asked to perform: segmentation, regression or classification. Natural Language Processing (NLP) is an application of AI algorithms that is recently gaining interest in the clinical domain.

**Figure 1:**
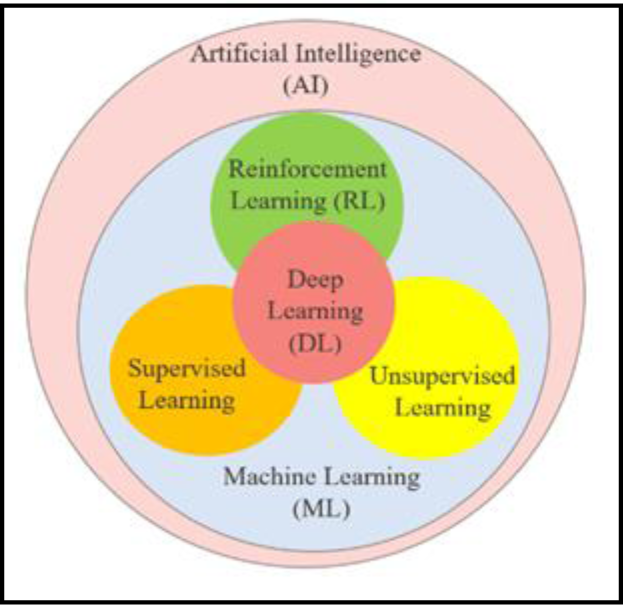
Types of Artificial Intelligence (AI) models.

AI/ML applications have begun to make inroads into the field of cancer pain prediction and management. These technologies offer the promise of more accurate pain assessment, personalized treatment recommendations, and improved patient outcomes [16]. However, the extent of their utilization and their impact on cancer pain prediction and management is an area that requires further investigation, and the results of the best performing models in cancer induced pain remain controversial and disperse.

In the dynamic landscape of AI/ML, the Transparent Reporting of a multivariable prediction model for Individual Prognosis or Diagnosis (TRIPOD) guidelines stand as a crucial framework for ensuring the transparency and reproducibility of predictive models. Developed to enhance the reporting quality of studies involving prediction models, TRIPOD provides a structured approach to the design, analysis, and interpretation of such models [17]. Adhering to these guidelines is paramount as it not only facilitates the effective communication of research findings but also fosters trust in the outcomes of predictive models. In the realm of ML, where complex algorithms increasingly influence decision-making across various health domains, the importance of adhering to TRIPOD guidelines is accentuated by the critical need for model validation [18]. Lastly, the validation process plays a pivotal role in assessing a model’s generalizability and reliability, ensuring that its predictive capabilities extend beyond the training data [17, 18].

The performance and generalizability of an AI/ML prediction model is assessed by testing the model on a data subset that has not been used during the model training process. The validation of a model can be internal or external, depending on whether the test dataset belongs to the same study cohort to that of the training dataset or is obtained from an entirely independent cohort. Both validation steps are crucial for the clinical implementation of a prediction model [18]. In particular, internal cross validation is a method to partition the data into training and testing subsets multiple times in order to provide a more accurate measure of model performance, especially with small datasets.

Comprehensive ML model performance assessment should include reporting of the model’s discrimination ability and model calibration as part of the internal and external validation process [19, 20]. Model discrimination is a measure of the model’s ability to differentiate between two classes (e.g., event vs. no event) given the available inputs [20]. Model calibration involves ensuring that the predicted probabilities align closely with the actual probabilities of events occurring [19].

Our scientific questions are:

- Which AI/ML algorithms are used in cancer pain research and cancer pain management?
- What are the applications of AI/ML models in cancer pain medicine?
- Which are the best performing models for cancer pain prediction and opioids optimization?
- To what degree do the existing AI/ML models in cancer pain prediction follow the TRIPOD guidelines with respect to model performance reporting?

This systematic review aims to answer these questions and address the current gap in knowledge by analyzing existing literature on the role of AI/ML in cancer pain prediction and management decision-making. We also provide an overview of common supervised and unsupervised learning techniques, their general characteristics, and specific applications in cancer pain research, either in cancer pain prediction, cancer treatment related pain or pain management and opioids decisions.

## Materials and Methods

### Protocol Registration

Registration of this systematic review in the international prospective register database of systematic reviews (PROSPERO), was done on 16 October 2023 [ID number: CRD42023469865] in the context of human health care.

### Search Strategy and Study Eligibility

We conducted a systematic search of Ovid MEDLINE, Ovid EMBASE, and Clarivate Analytics Web of Science, for publications in English from the inception of databases to September 7, 2023. The concepts searched included “Cancer”, “Pain”, “Pain Management”, “Pain Measurement”, “Analgesics”, “Opioids”, “Artificial Intelligence”, “Machine Learning”, “Deep Learning”, “Expert System” and “Neural Networks”. Both subject headings and keywords were utilized. The terms were combined using AND/OR Boolean Operators. Animal studies, in vitro studies, and conference abstracts were excluded. The complete search strategy is detailed in Tables S1–S3.

### Screening process

Identified articles from the Search process were uploaded into the Covidence screening application [21], and screening through Covidence was conducted by two independent reviewers. Screening of the titles and abstract was first conducted and then screening of the full text was done on the retrieved articles. Final included articles were extracted from Covidence for full-text review and data collection.

### Inclusion Criteria

Studies eligible for inclusion in this systematic review had the following criteria 1) be published in English, 2) investigate AI/ML applications in cancer pain medicine or cancer pain management, and 3) involve human subjects.

### Exclusion Criteria

Articles were excluded if they met any of the following criteria: 1) out of scope of our study 2) no AI/ML application, 3) non-cancer pain study, 3) not an original study (i.e., review article, letter, conference abstract), 4) duplicate publication or correction of an original article, 5) descriptive study that didn’t apply or test models.

### Data Synthesis

This study followed the Preferred Reporting Items for Systematic Reviews and Meta-Analysis (PRISMA) guidelines [22].

### Data Collection Process

The collected data included the following: AI/ML technique(s) used, input features and output variables, validation methods, and model performance metrics. Adherence to TRIPOD guidelines was analyzed using TRIPOD checklist [18]. Included articles were categorized according to the use of the AI/ML in cancer pain and grouped into the following groups: (1) models for prediction of cancer related pain intensity, cancer pain diagnosis or cancer pain initiation, (2) models for prediction of post cancer treatment pain (e.g., cancer surgery, RT or CT), (3) models for cancer pain management prediction or as a decision support system for cancer pain management.

## Results

### Search and screening Results

This comprehensive search resulted in identification of 436 studies through search of MEDLINE (n=189), EMBASE (n=189) and Web of Science (n=132). After screening and eligibility assessment, 283 articles were excluded. A total of 44 studies met the inclusion criteria and were included in this review. The search strategy process is illustrated in a PRISMA flow diagram (Figure 2). All included studies were published between 2006 and 2023, with an increase in the number of publications for AI/ML studies in cancer pain research from 1 (2006-2009) to 26 (2020-2023) (Figure 3).

**Figure 2:**
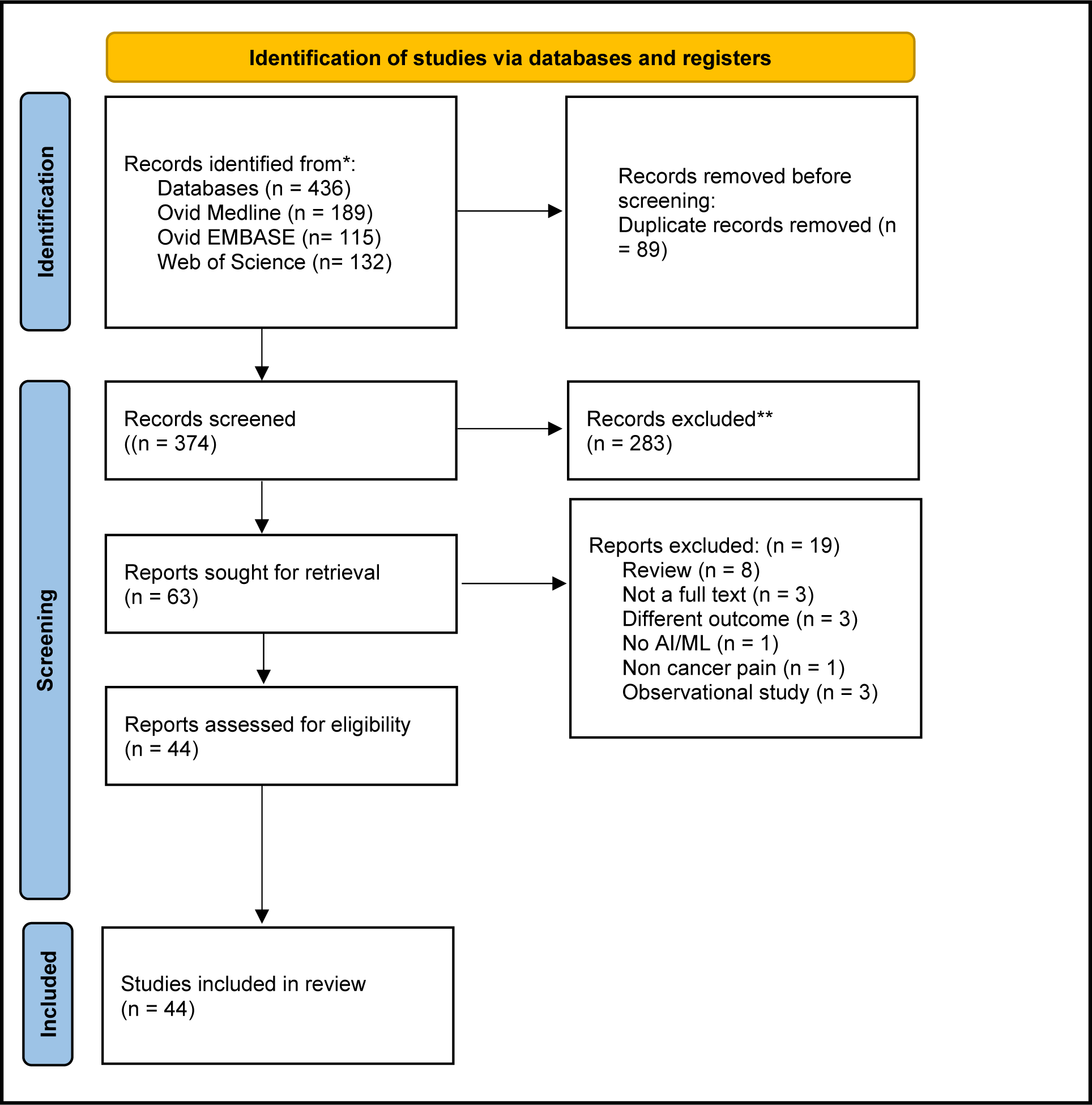
PRISMA Flow Diagram for systematic reviews of AI and ML in cancer pain res.

**Figure 3:**
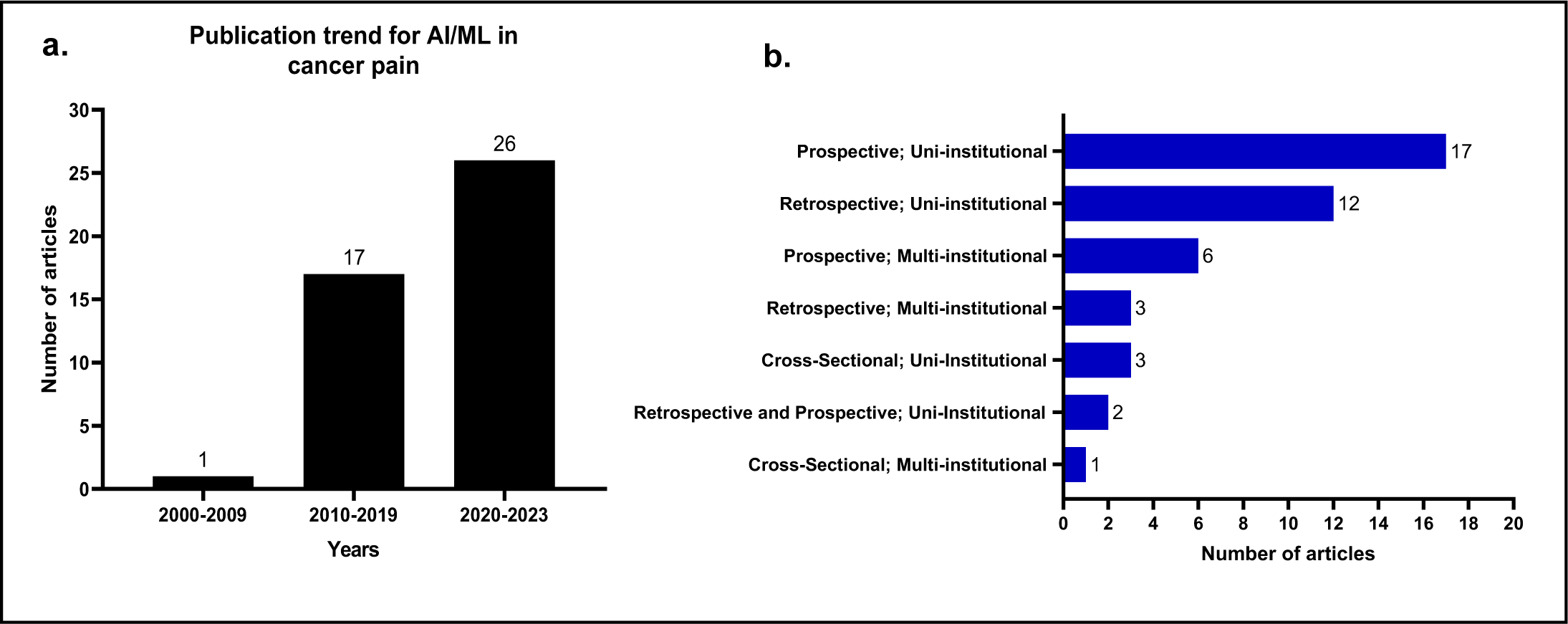
**a.** Publications trends for AI/ML models used for cancer pain research between 2006-2023. **b.** Types of studies and the number of articles per each type.

### Design and populations

Most studies used a prospective uni-institutional cohort to develop their models (n=17 studies) while the least used approach involved the cross-sectional multi-institutional cohort design (n=1) (Figure 3.b). The median sample size to build the models was 320 (range: 21-46104, IQR 140-1000) 95% CI 156-900. Five studies did not specify the size of the study population. In 95% (n=23) of studies the cohort size was between 100-1000 patients, while 23% (n=9) of studies used >1000 patients and 18% (n=7) used <100 patients.

### AI/ML algorithms used in cancer pain research

The most common AI/ML algorithms identified in this review, their general characteristics and specific use in cancer pain research are summarized in (Table 1). Some studies used a single model, while the majority of studies explored multiple models (each model was explored separately) (55%, n=24 articles) (Figure 4.a). The most common algorithms used in these multiple models studies were Random Forest (RF) and Logistic Regression (LR) (n=15 studies for each) (Figure 4.b). Other single model studies included Neural Networks (NN) (16% n=7), Decision Trees (DT) (7% n=3), Decision support system-Fuzzy (n=2), Clustering (n=2), LR (n=2), Bayesian networks (n=1), NLP (n=1) and 2 studies used novel models (Figure 4.a). Several other models used in multiple models studies (e.g., SVM, transformer, RF, GBM, Lasso, and NB) (Figure 4.b)

**Table 1:** AI/ML model characteristics and applications in cancer pain research.

| Machine Learning Algorithm | Characteristics | Cancer Pain Usage |
| --- | --- | --- |
| **<u>Supervised Learning</u>**<br>[23] | The output is labeled with the desired value (e.g., pain score or opioids dose) |  |
| **Classification** | The output variable is a binary and/or categorical response |  |
| Decision trees (DT)<br>[24], [25], [26], [27], [28], [29], [16], [30], [31] | Generates understandable rules with both categorical and continuous variables for prediction and classification purposes. | • Cancer pain prediction<br>• Post cancer treatment pain prediction<br>• Cancer pain analgesics requirement |
| Random Forest (RF)<br>[24], [32], [33], [34], [26], [35], [27], [36], [37], [16], [10], [38], [39], [40], [41] | An ensemble approach that combines the output of multiple decision trees to reach a single high accurate result. It provides a good predictive performance, low overfitting, and easy interpretability. | • Cancer pain prediction<br>• Post cancer treatment pain prediction<br>• Cancer pain analgesics requirement<br>• Cancer pain related unscheduled healthcare prediction<br>• Prediction of remote consultation/visits or patient satisfaction due to cancer pain. |
| Gradient boosting (GBM); extreme gradient boosting (XGBoost)<br>[32], [33], [35], [27], [28], [37], [16], [38], [41] | A robust ensemble boosting algorithm that trains the model sequentially for prediction and classification purposes. It combines several weak learners into strong learners. | • Post cancer treatment pain prediction<br>• Cancer pain related unscheduled healthcare prediction<br>• Cancer pain analgesics requirement<br>• Cancer persistent pain prediction |
| Naïve Bayes (NB) Classifier<br>[26], [37], [16] | Used for binary or multi-class classification problems, based on Bayes rule and conditional independence assumption. | • Post cancer treatment pain prediction<br>• Cancer persistent pain prediction |
| **Regression** | Predicts the probability using continuous data. |  |
| Generalized linear mixed models (GLMMs), least absolute shrinkage and selection operator (Lasso), Linear regression (LR), Logistic regression (LR), Bayesian regression [42], [32], [43], [33], [34], [35], [27], [44], [45], [46], [39], [41], [31] | Model that estimates the relationship between one dependent variable and one or more independent variables using a line. | • Cancer pain prediction<br>• Post cancer treatment pain prediction<br>• Cancer pain analgesics requirement<br>• Cancer pain related unscheduled health care prediction<br>• Association between epigenetics and persistent cancer pain |
| **Support vector machine (SVM)**<br>[43], [34], [47], [26], [27], [28], [37], [40], [41] | Type of supervised learning algorithm used to solve classification and regression tasks. Creates a hyperplane to separate two classes. | • Cancer pain prediction<br>• Post cancer treatment pain prediction<br>• Cancer pain analgesics requirement |
| **<u>Unsupervised Learning</u>**<br>[36] | Unlabeled and unclassified datasets used to train machines; Used to categorize unsorted data based on features, similarities and differences. | • Mapping parameters and post cancer treatment persistent pain |
| **Clustering**<br>[25], [48], [49] | Machines divide the data into clusters based on features, similarities and differences. | • Risk features identification for cancer pain<br>• Identifying groups sharing patterns in post cancer treatment persistent pain |
| **Association**<br>[50] | Machines find interesting relations and connections among variables within large datasets that are input. | • Correlation between genetic SNPs and post treatment cancer pain outcome. |
| **Neural Networks (NN)**<br>[33], [25], [34], [26], [27], [28], [51], [52], [16], [10], [46], [38], [53], [54], [55], [56], [31] | Composed of node layers, containing an input layer, one or multiple hidden layers and an output layer. Could be supervised, unsupervised or semi-supervised. | • Cancer pain prediction<br>• Post cancer treatment pain prediction<br>• Cancer pain identification using video analysis<br>• Prediction of remote consultation/visits due to cancer pain.<br>• Association between epigenetics and persistent cancer pain |
| Deep Learning (DL)<br>[57], [56] | Subclass of NN that Includes many layers of the neural network and massive volumes of complex and disparate data. | Genetic mutation associated cancer pain<br>Cancer pain prediction |
| **<u>Natural Language Processing (NLP)</u>**<br>[58], [59], [60] | A component of AI that uses the ability of a computer program to understand human language either written or spoken, it could be supervised or unsupervised. | Identification of cancer pain and cancer attributes |
| **<u>Decision Support System</u>**<br>[61], [62], [31] | A subtype of AI using computerized programs to help decision making, judgment and actions in an organization. | Evaluate decision support computer program for cancer pain management.<br>Pain treatment recommendations and staging<br>Pain intensity identification |

**Figure 4:**
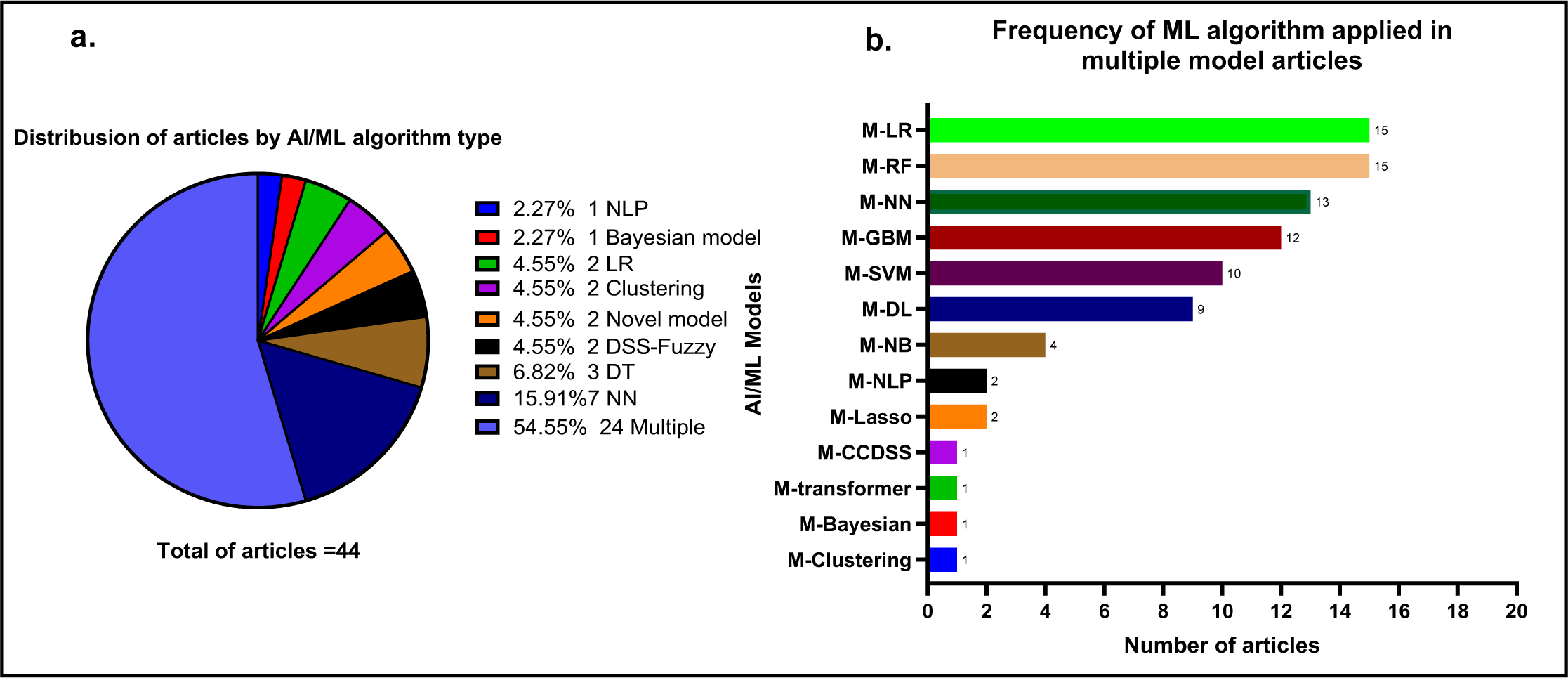
**a.** Distribution of articles by AI/ML algorithm type. **b.** ‘Frequency of ML algorithm applied in multiple model articles’ (*M; multiple*)

### AI/ML Models used for cancer related pain research

This review identified 18 studies describing the implementation of AI/ML in cancer pain research (Table 2).

**Table 2:**
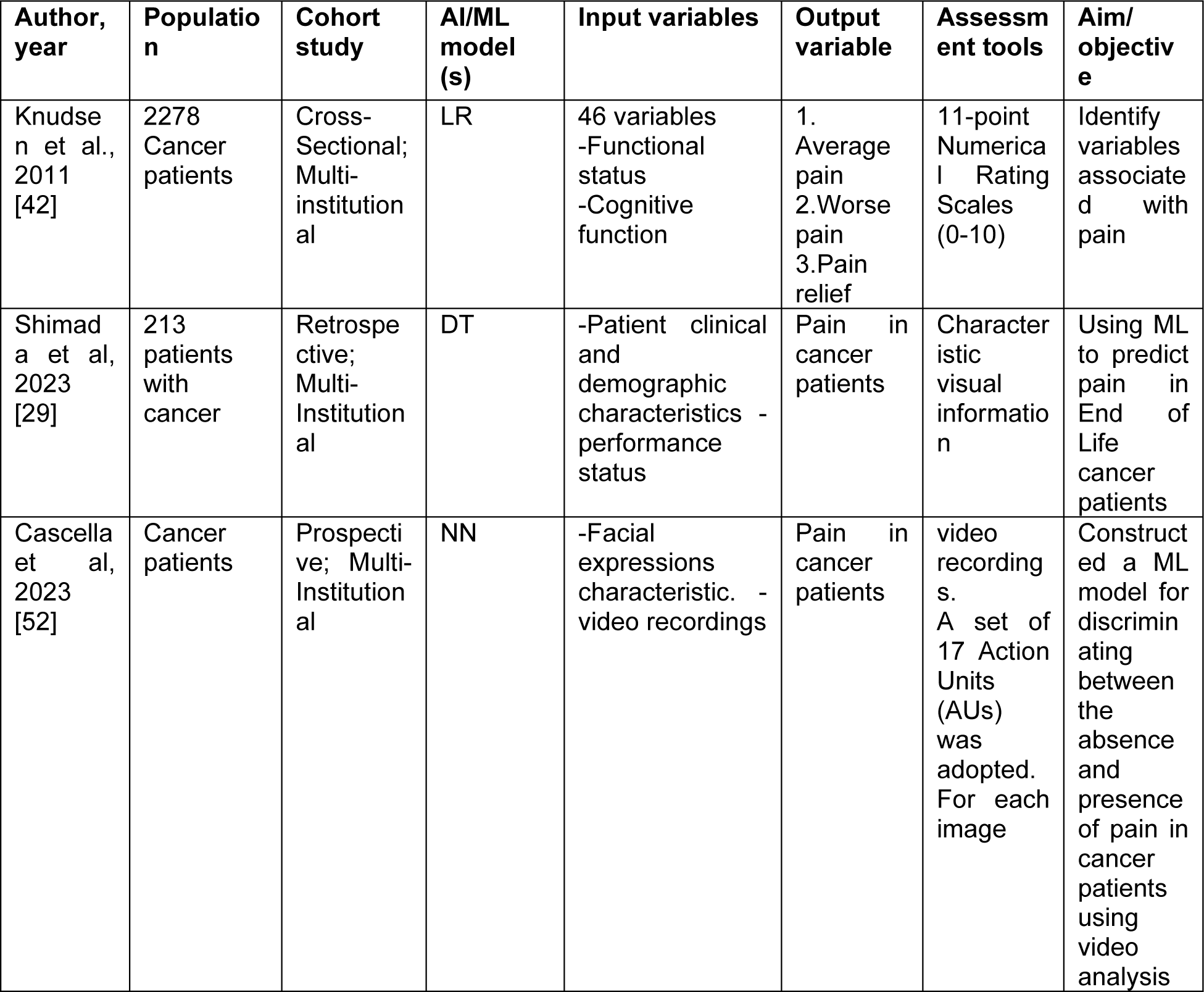
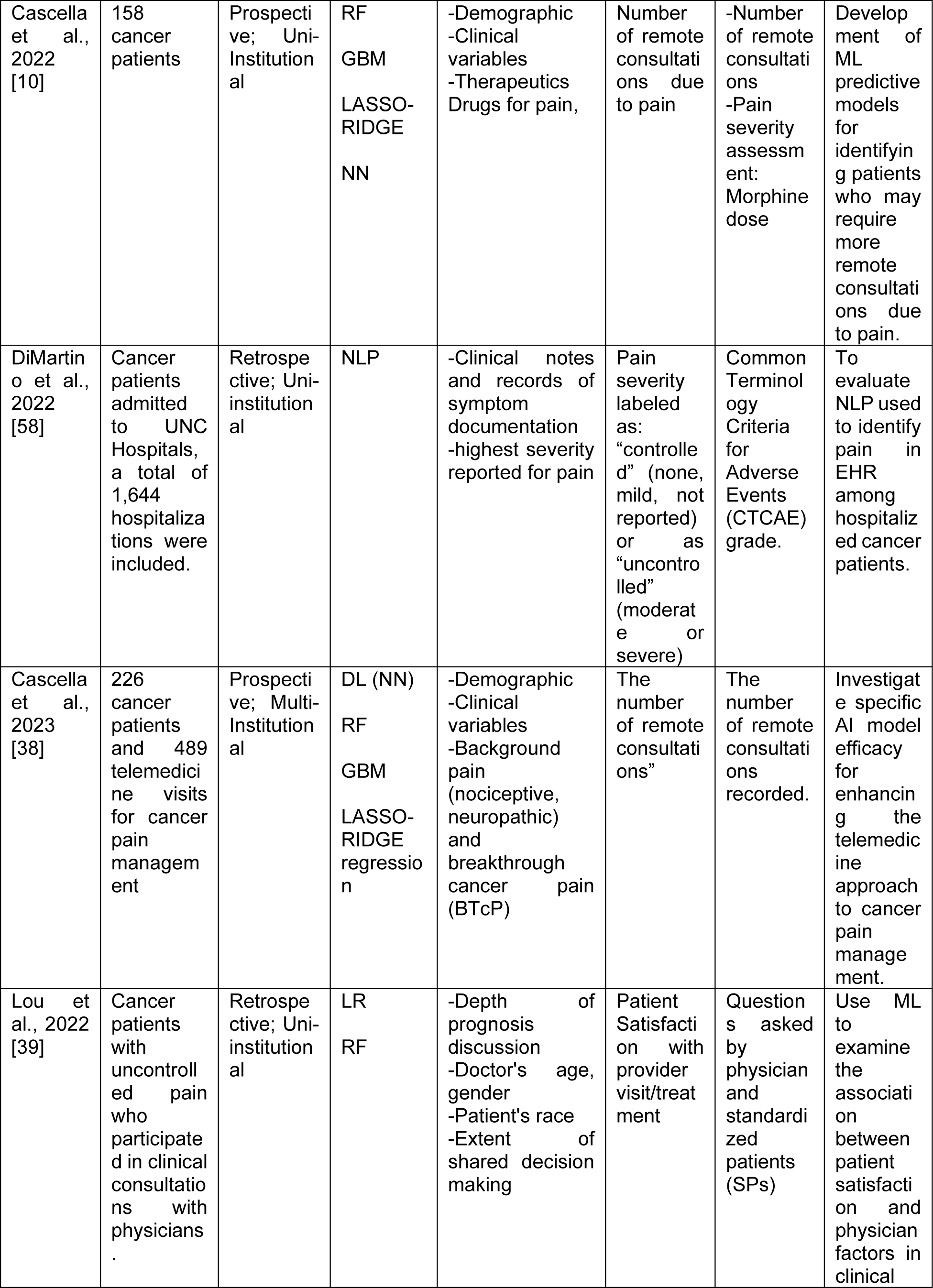
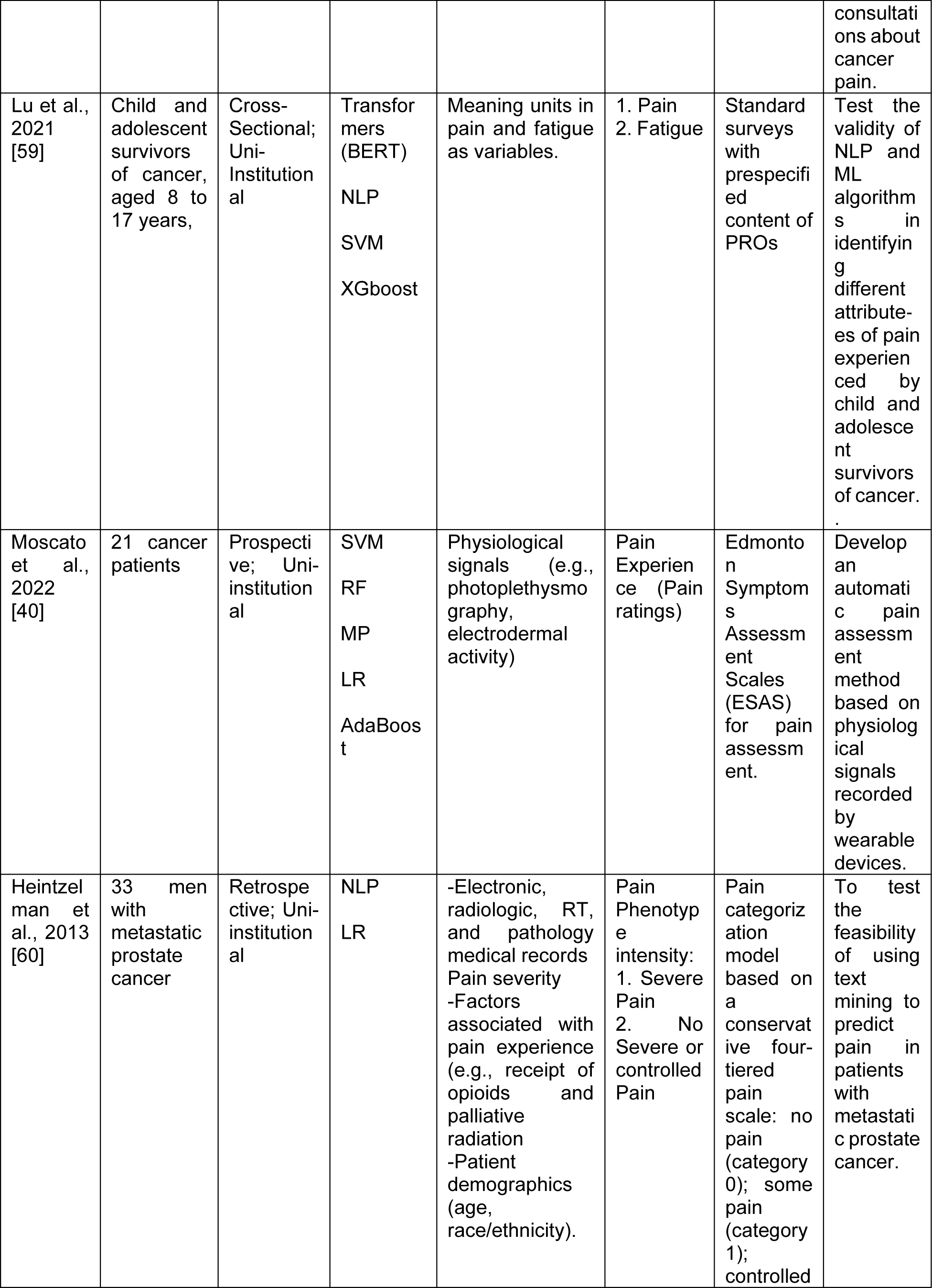
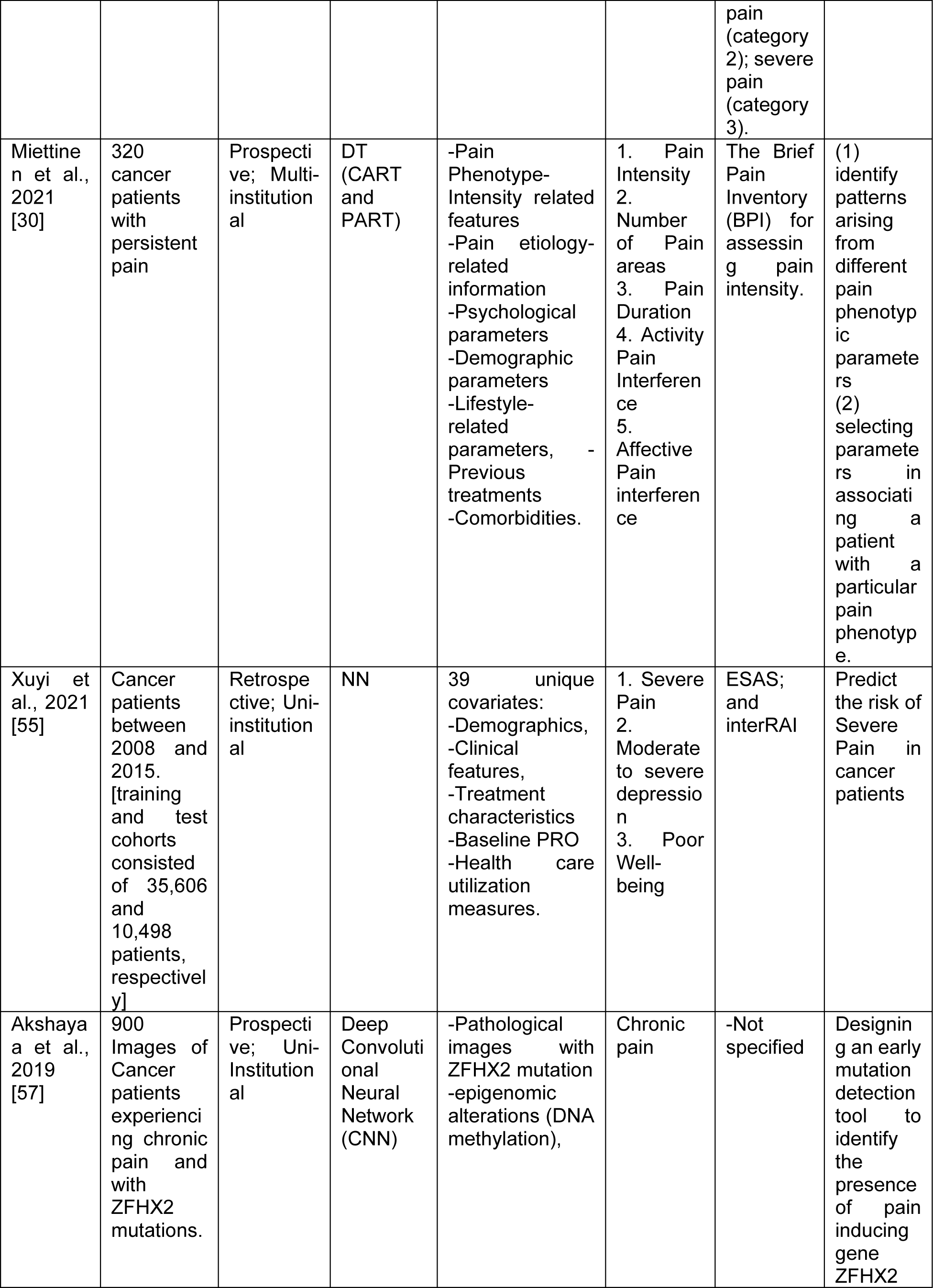
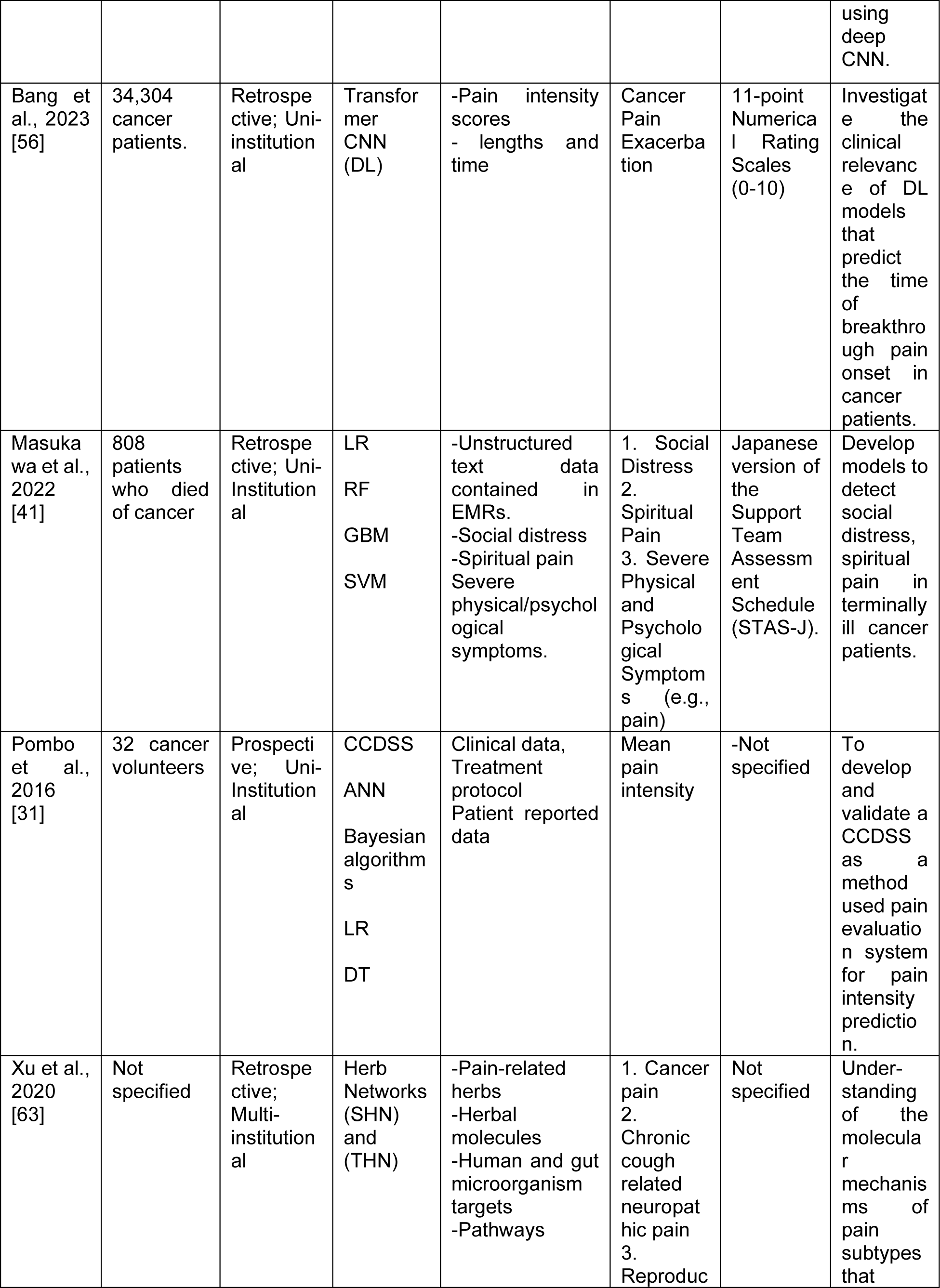
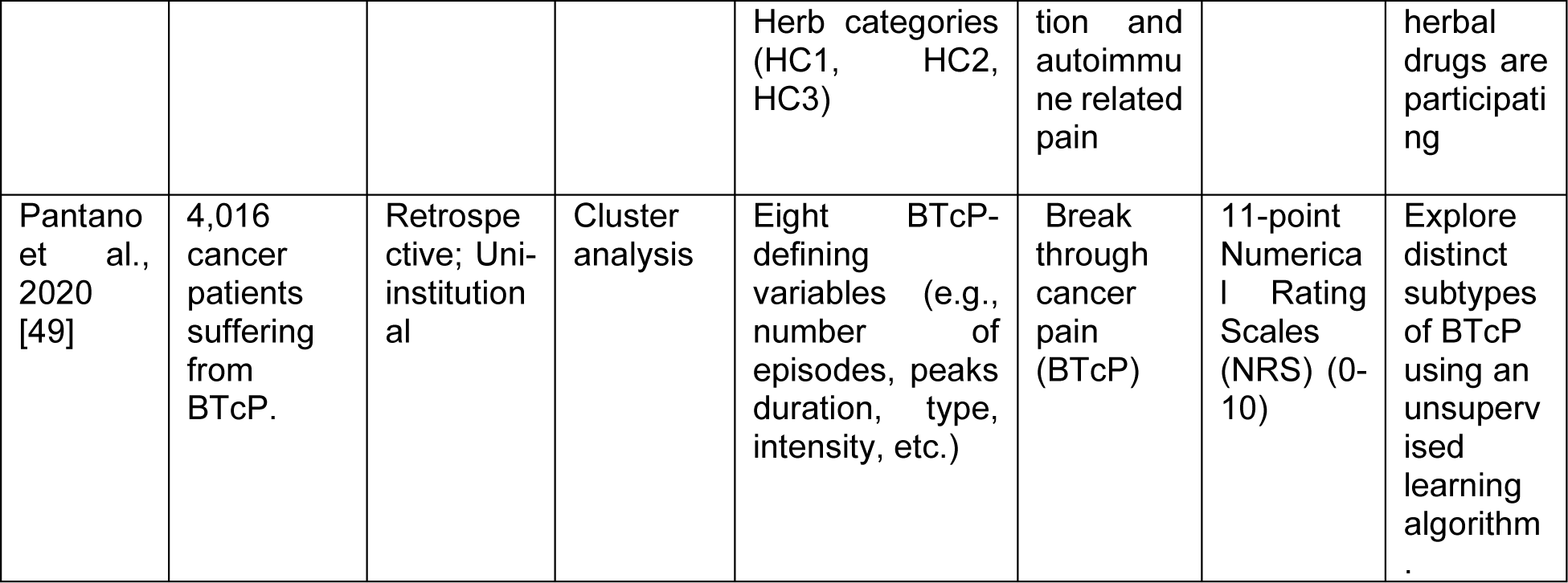
Characteristics of studies that used AI/ML algorithms in cancer pain research.

In a study by Knudsen et al. (2011) [42] a cross-sectional analysis of 2278 cancer patients utilized Linear regression to identify key variables associated with pain, revealing breakthrough pain, psychological distress, sleep issues, and opioid dose as significant factors. Shimada et al. (2023) [29] employed DT in a retrospective study predicting end-of-life cancer patients’ pain, showcasing the potential of ML in understanding complex patient scenarios. Cascella et al. (2023) [52] and Cascella et al. (2022) [10] explored video analysis and telemedicine, respectively, employing NN and various models to enhance cancer pain management. DiMartino et al. (2022) [58] used NLP to evaluate uncontrolled symptoms (i.e., pain), showing promise of AI in pain prediction (accuracy 61%). Lou et al. (2022) [39] delved into patient satisfaction in cancer pain consultation, using LR and RF, revealing associations with physician factors but facing limitations in data collection and ML metric reporting. Lu et al. (2021) [59] tested ML algorithms on young cancer survivors, with Bidirectional Encoder Representations from Transformers (BERT) showing higher accuracy in identifying pain interference. Moscato et al. (2022) [40] explored physiological signals for pain assessment. Results demonstrated the feasibility of using ML for pain assessment via physiological signals in real-world contexts. Heintzelman et al. (2013) [60] demonstrated the feasibility of text mining using NLP for predicting pain severity in metastatic prostate cancer. Miettinen et al. (2021) [30] identified pain phenotype clusters using DTs, while Xuyi et al. (2021) [55] predicted multiple symptoms using NNs. Akshayaa et al. (2019) [57] achieved high accuracy in detecting chronic pain with CNN, and Bang et al. (2023) [56] predicted breakthrough pain onset with DL models. Masukawa et al. (2022) [41] utilized LR, RF, GBM, and SVM to detect social distress and spiritual pain in palliative care. Pombo et al. (2016) developed a Computerized clinical decision support system (CCDSS) for pain intensity prediction, showing accuracy but limited clinical validation discussion. Xu et al. (2020) [63] explored herbal drugs’ molecular mechanisms in pain subtypes. Pantano et al. (2020) [49] identified Break through cancer pain (BTcP) clusters.

These studies employ various AI/ML models, showcasing their potential in understanding patient satisfaction, identifying pain-related attributes, and predicting pain in cancer patients across different age groups and healthcare settings. Most studies used clinical and demographic variables as input; however, some studies applied models utilizing innovative inputs such as physiological signals [40], facial expressions [52], textual data [41], and even herbal categories [63], indicating the exploration of novel data sources. This diversity in inputs strengthens the versatility of the models. Some studies showcase innovation by assessing pain through video analysis of facial expressions [52], wearable devices capturing physiological signals [40], and even employing NLP for identifying pain severity in unstructured medical records [58]. These novel approaches strengthen the potential for non-traditional but effective pain assessment.

Most studies lacked detailed information on the extent of external validation of the AI models applied and the clinical application in real healthcare scenarios. Only 3 out of 18 studies performed external validation, and 7 studies discussed the clinical evaluation and application of the models used. Cascella et al, 2022 & 2023 [10] [38] applied external validation and clinical evaluation of the models for the remote consultation prediction in cancer patients. Additionally, Cascella et al, 2023 [52] performed a clinical trial to test the NN using video recording of facial expression for discriminating between the absence and presence of pain in cancer patients. Heintzelman et al., 2013 [60] proved the feasibility and generalizability of NLP through an external validation using separate data source, furthermore, they evaluated the clinical application of the NLP using text mining for pain prediction in patients with metastatic prostate cancer. Pombo et al., 2016 [31] clinically evaluated the CCDSS compared with clinical advice for pain intensity, however, there was a lack of generalizability of the decision model due to the uni-institutial cohort. Moscato et al., 2022 [40] applied a real-world context to develop an automatic pain assessment model, however, the small size cohort was a limitation with the need for bigger cohort for clinical validation.

### AI/ML Models used for cancer treatment induced pain research

Nineteen articles focused on implementing AI/ML algorithms in cancer treatment pain research were detected in our review. The characteristics of these studies are summarized in Table 3.

**Table 3:**
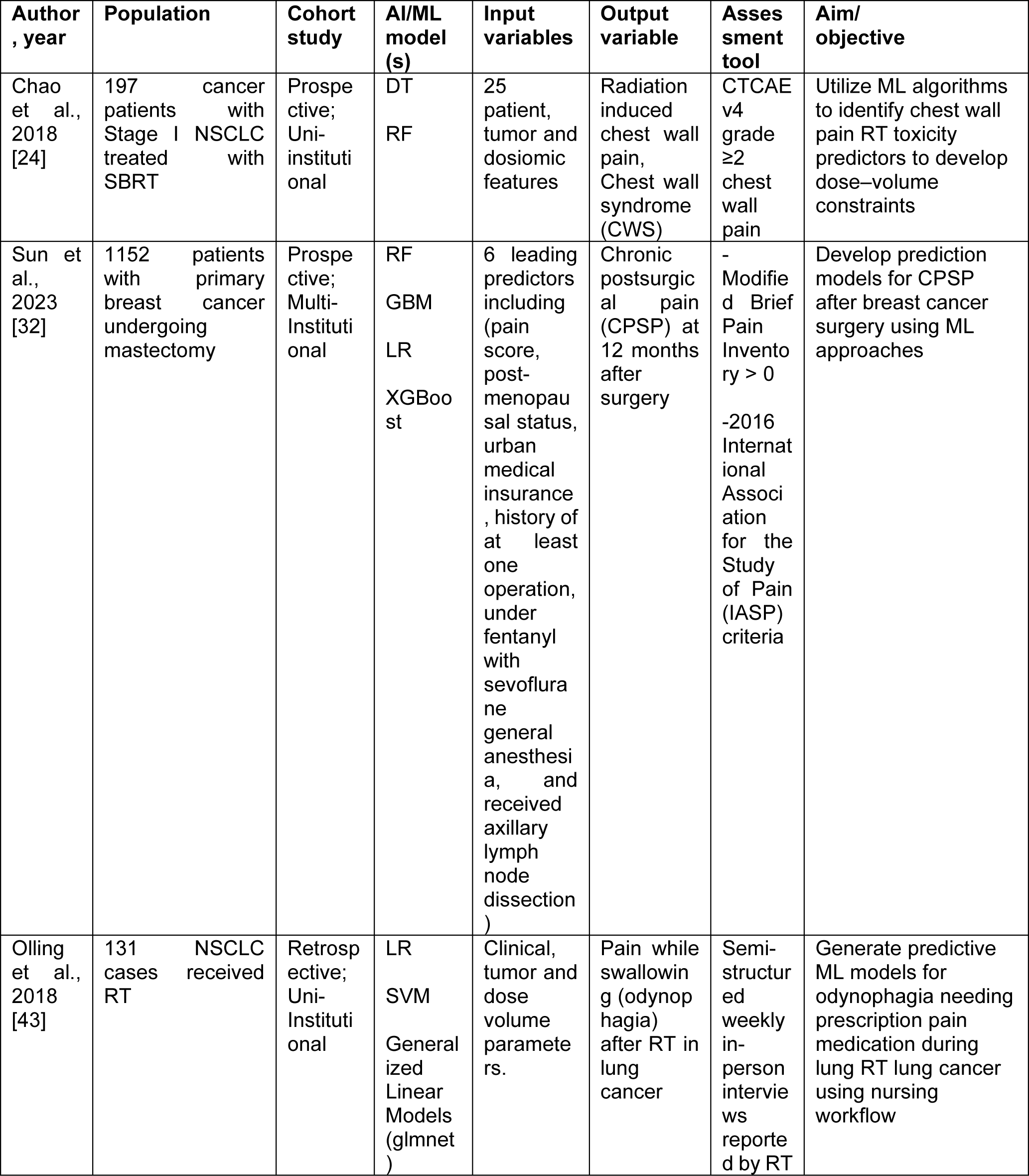
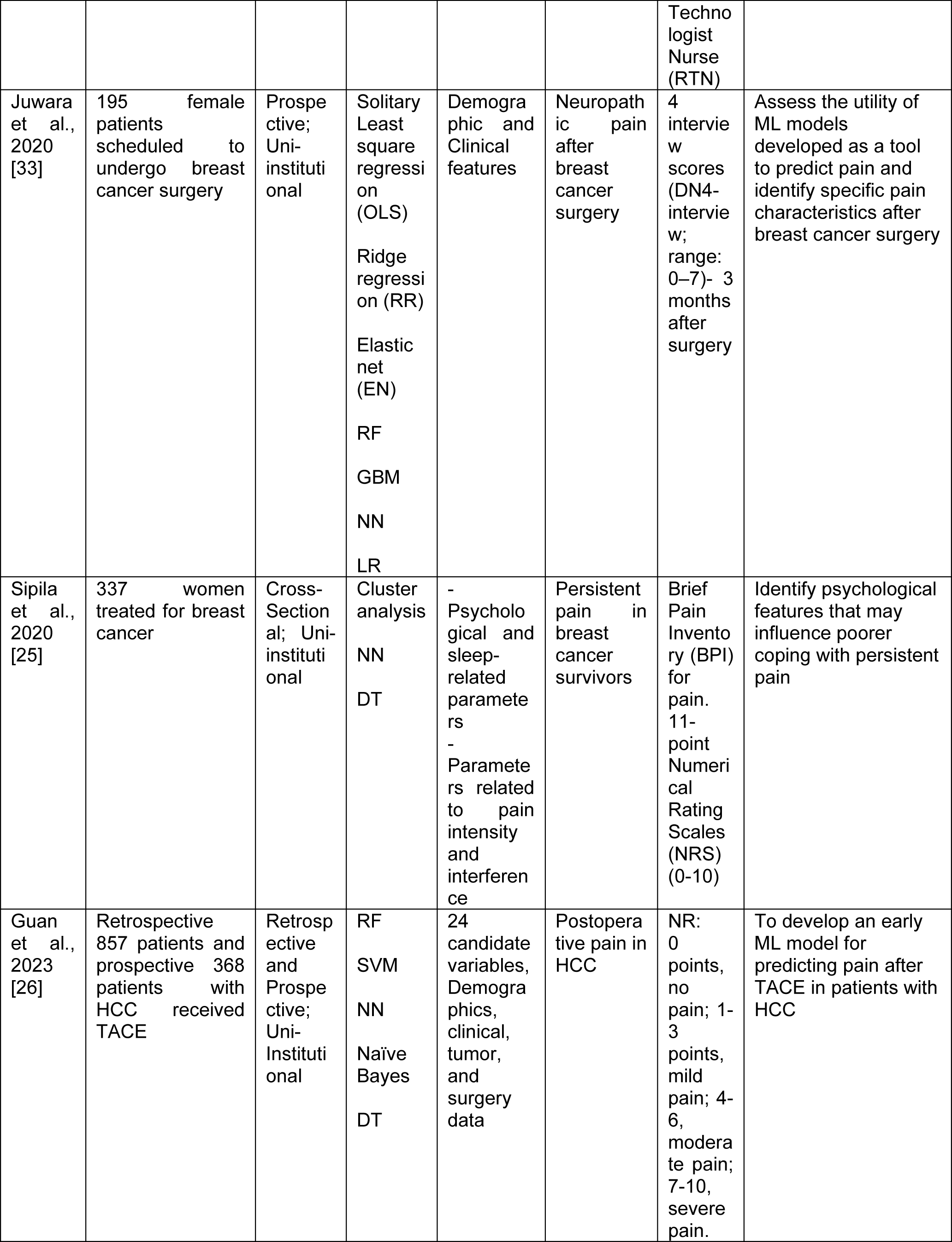
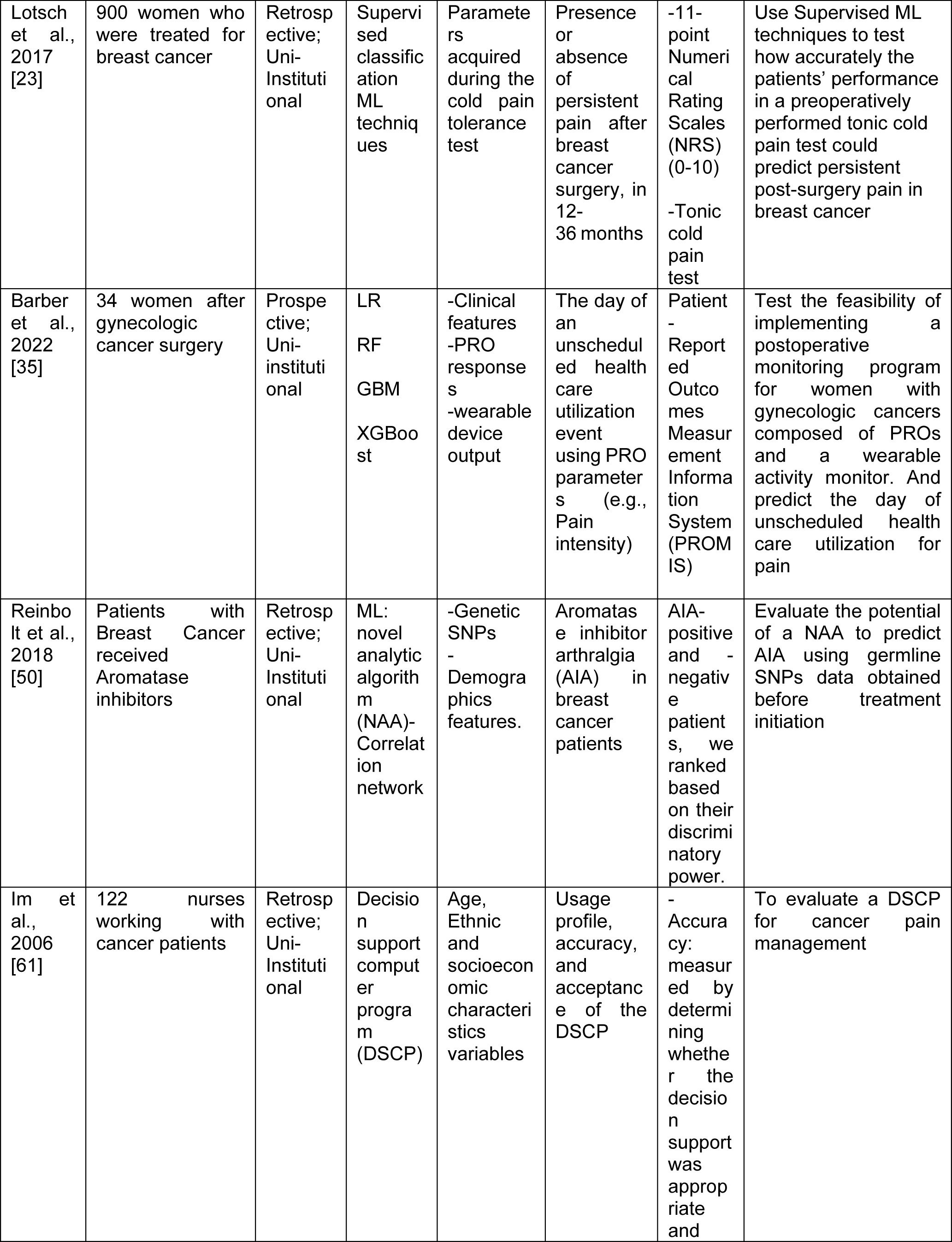
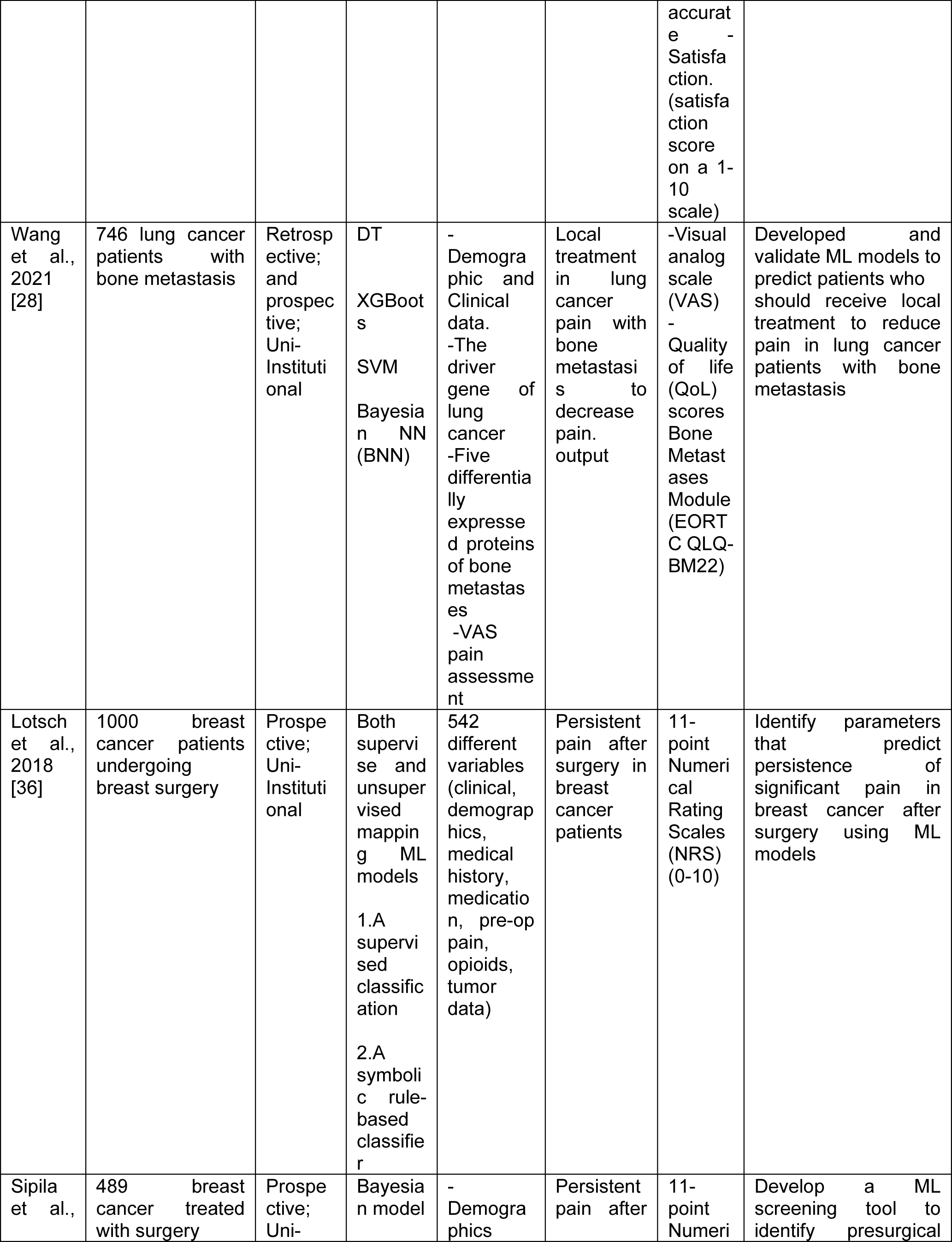
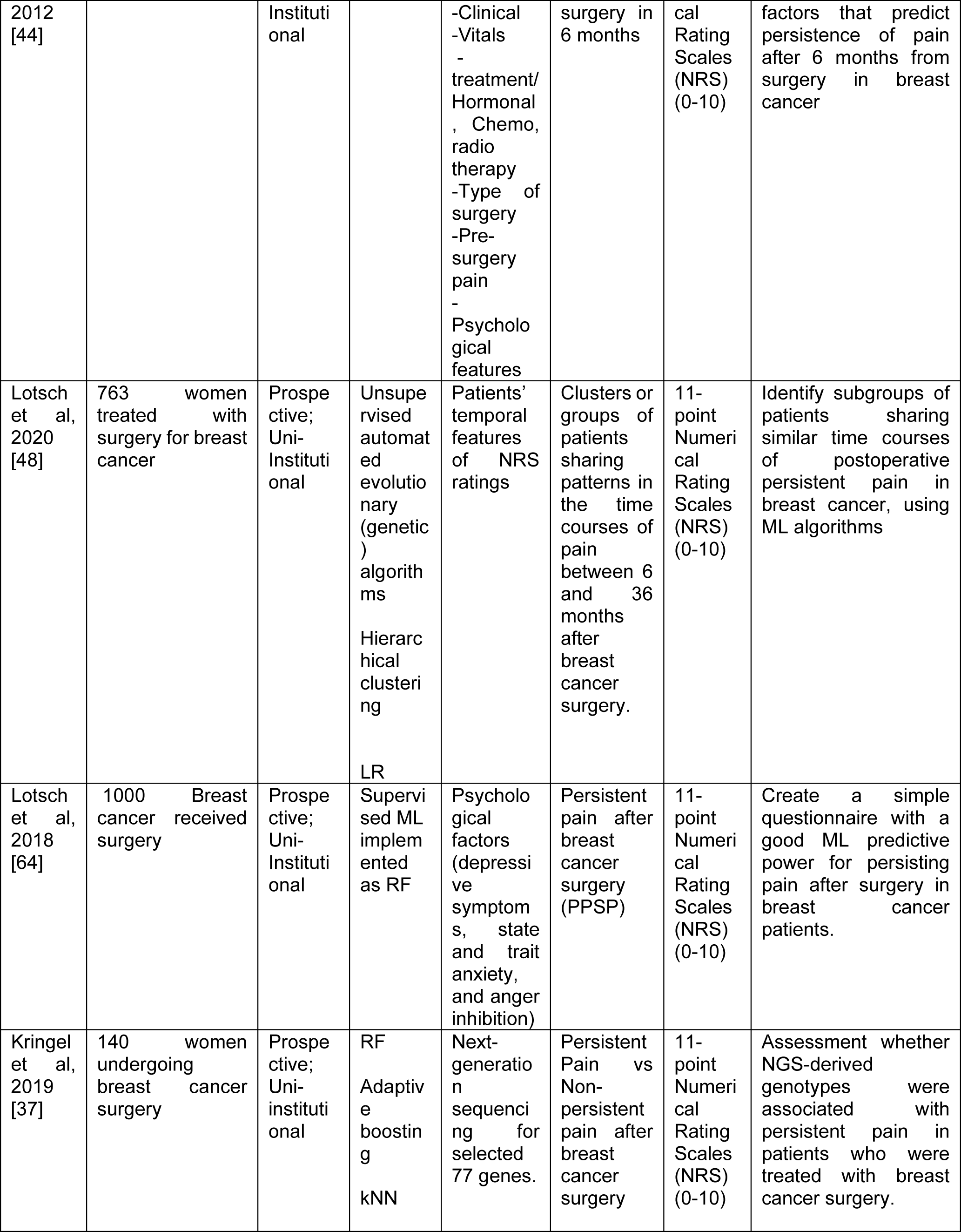
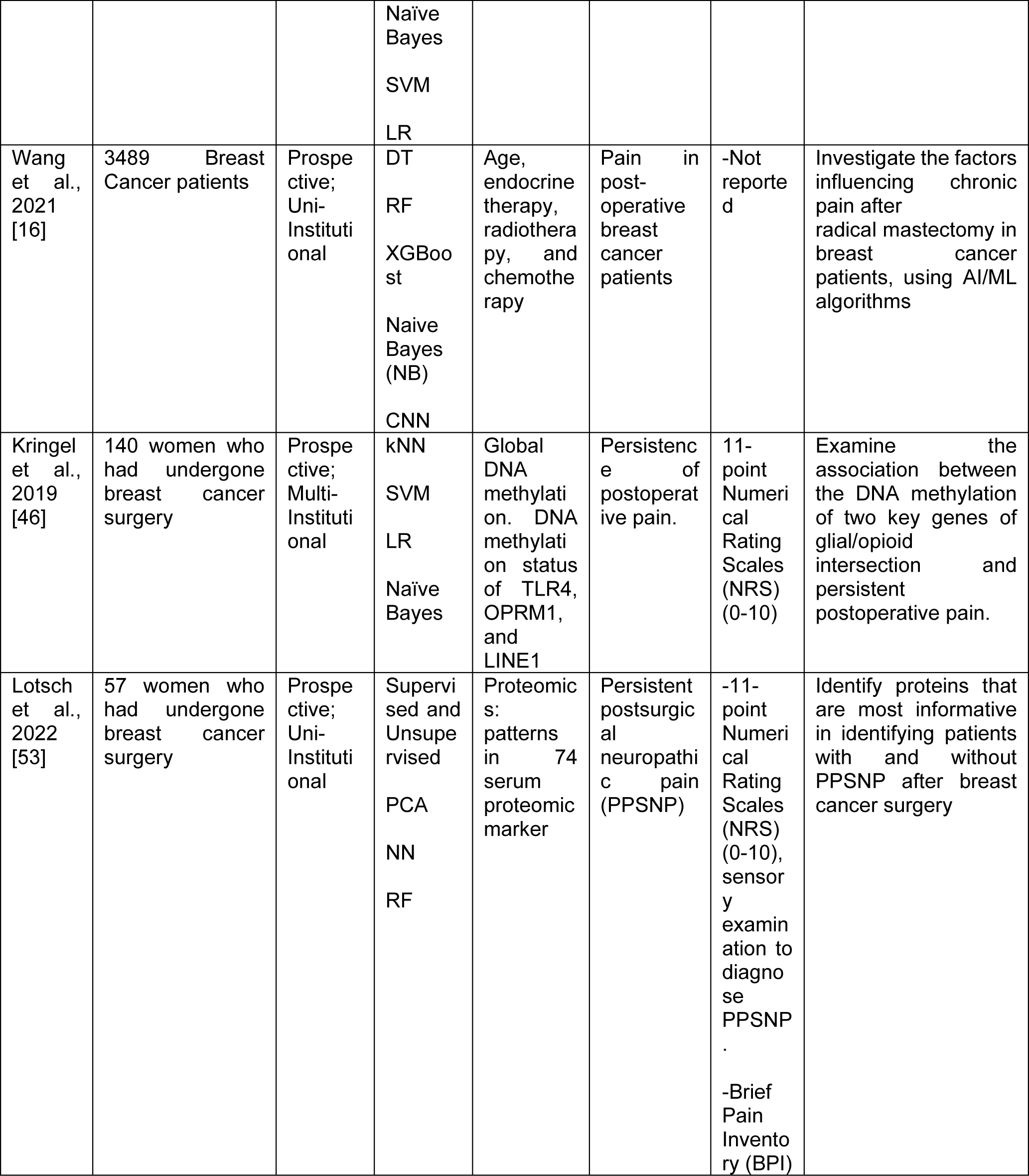
Characteristics of studies used AI/ML algorithms in cancer treatment related pain research.

The study by Chao et al. in 2018 [24] aimed to predict radiation-induced chest wall pain in Non-Small Cell Lung Cancer (NSCLC) patients using DT and RF models. The study demonstrated that ML models are predictive for chest wall pain toxicity after RT in Lung cancer patients. Additionally, Olling et al. (2018) [43] conducted a study aimed at predicting pain while swallowing (odynophagia) post-RT in lung cancer, applying LR, SVM and Generalized Linear Models (glmnet) based on various parameters. These models accurately predicted odynophagia during lung cancer RT, revealing their effectiveness in pain prediction during the course of treatment for lung cancers. Studies conducted by Sun et al. (2023) [32], Juwara et al. (2020) [33], Lotsch et al. (2017) [23], Lotsch et al. (2018) [36], Sipila et al. (2012) [44], Lotsch et al. (2018) [34] and Wang et al. (2021) [27], investigated various ML models for persistent/chronic pain prediction and features identification in post-surgery in breast cancer patients. Studies by Sun et al. (2023) [32] and Juwara et al. (2020) [33] revealed that the novel ML approaches, including RF, GBM, and XGBoost, exhibited higher performance in predicting Chronic Postsurgical Pain (CPSP) over the traditional regression models. Lotsch et al. (2017) [23] revealed that supervised classification ML techniques robustly excluded post-surgical persistent pain, exhibiting a high accuracy of 94.4% based on preoperative cold pain sensitivity. Sipila et al. (2012) [44] developed a screening tool using ML to identify risk factors contributing to persistent post-surgery pain using a Bayesian model. The study’s ML model showed high predictive performance and identified significant predictors of prolonged pain after breast cancer surgery, such as preoperative chronic pain and multiple previous operations. Lotsch et al. (2020) [48] used automated evolutionary algorithms, hierarchical clustering, and LR to identify distinct pain patterns from 6 to 36 months post-surgery and revealed three unique patient groups exhibiting differing persistent postoperative pain patterns. Wang et al. (2021) study results demonstrated that XGBoost algorithm exhibited the highest precision in predicting chronic pain, while the Naive Bayes (NB) model showed commendable performance in recall rate and F1 score. Sipila et al. (2020) [25] used various ML techniques to identify psychological factors influencing persistent post-treatment pain. They found that psychological and sleep-related parameters played a crucial role in grouping patients dealing with persistent pain after breast cancer treatments.

In a retrospective and prospective study, Guan et al. (2023) [26] investigated postoperative pain in hepatocellular carcinoma (HCC) patients after trans arterial chemoembolization (TACE). Using various ML models, they found that the RF model accurately predicted the risk of pain following TACE in HCC patients. Barber et al. (2022) [35] investigated a group of 34 women after gynecologic cancer surgery, utilizing ML models such as LR, RF, GBM, and XGBoost. They employed patient-reported outcomes (PROs) and wearable device data to predict the specific day of unscheduled healthcare utilization events, with a particular emphasis on pain intensity. The study’s primary finding was the feasibility of a PRO-based program in accurately predicting the occurrence of unscheduled healthcare utilization following surgery, especially concerning pain.

Reinbolt et al. (2018) [50] examined a novel analytic algorithm (NAA) to analyze genetic single nucleotide polymorphisms (SNPs) for predicting Aromatase Inhibitor Arthralgia (AIA) in breast cancer patients treated with Aromatase Inhibitors (AIs). Their key finding was the identification of 70 SNPs from 57 genes that predicted AIA with a notable accuracy of 75.93%. The study demonstrated the potential of using genetic markers to anticipate AIA, providing a promising avenue for preemptive risk identification. Furthermore, Kringel et al. (2019) [46] utilized kNN, SVM, LR, and NB, their ML analysis to understand the association between DNA methylation of glial/opioid-related genes and persistent post-surgery pain in breast cancer patients. Their study highlighted that global DNA methylation held a comparable diagnostic accuracy for persistent pain, compared to established non-genetic predictors. Finaly, Lotsch et al. (2022) [53] explore the potential of ML techniques such as (PCA, NN, RF) to identify proteins that distinguish patients with persistent postsurgical neuropathic pain (PPSNP). The study revealed 19 proteins that showed significant differences between the groups, marking a key finding.

As for the previous set of studies, most studies considered in this subgroup also lacked detailed information on the extent of external and clinical validation in real healthcare scenarios. Three out of 19 studies, used external clinical validation of the AI/ML models. Guan et al., 2023 and Wang et al., 2021 [26] [28] used an external prospective clinical cohort for clinical evaluation of the models. Additionally, Im et al., 2006 [61] used external cohorts of nurses from the internet for the clinical evaluation of the decision support computer program (DSCP) developed for pain prediction and management. Although studies used genomic, epigenomic and proteomic data to build the classification and clustering pain models, got the identified molecules from the clinical settings, were previously identified, however, no actual clinical validation and application were used, and the clinical evaluations of the models are still needed.

### AI/ML Models used for cancer pain management prediction and decision

Seven articles focused on implementing AI/ML algorithms in cancer pain management research were detected in our review. The characteristics of these studies are summarized in Table 4.

**Table 4:**
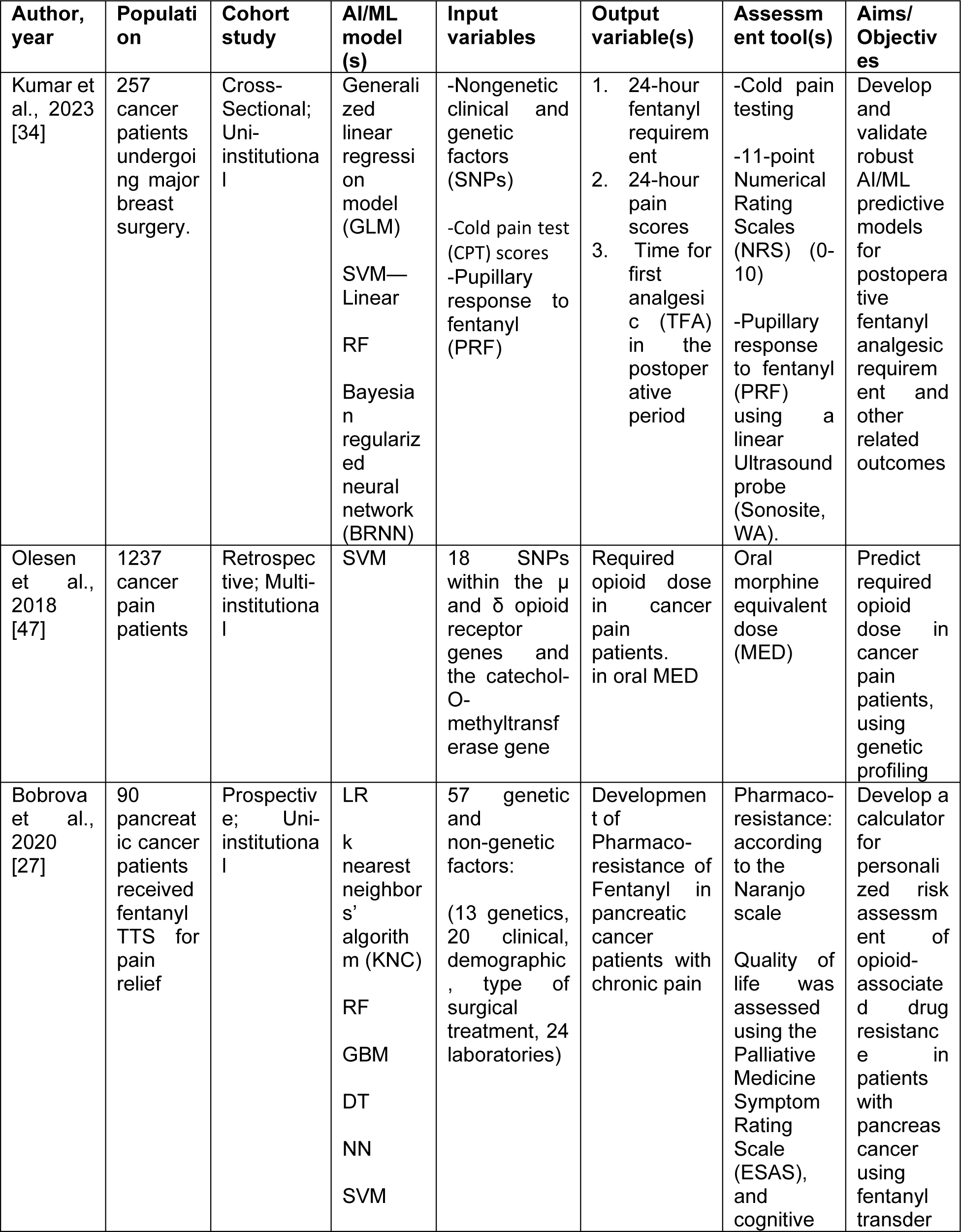
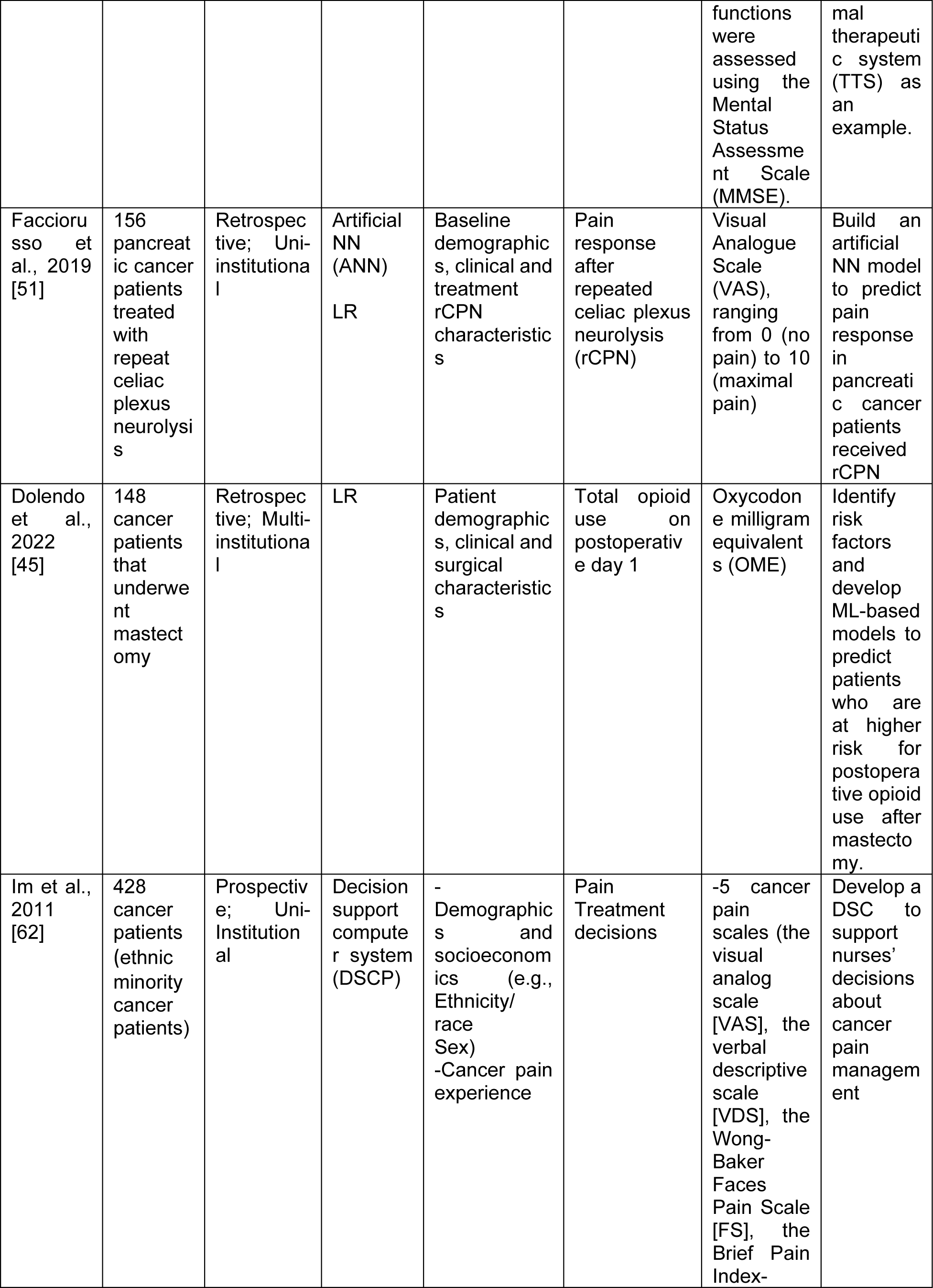
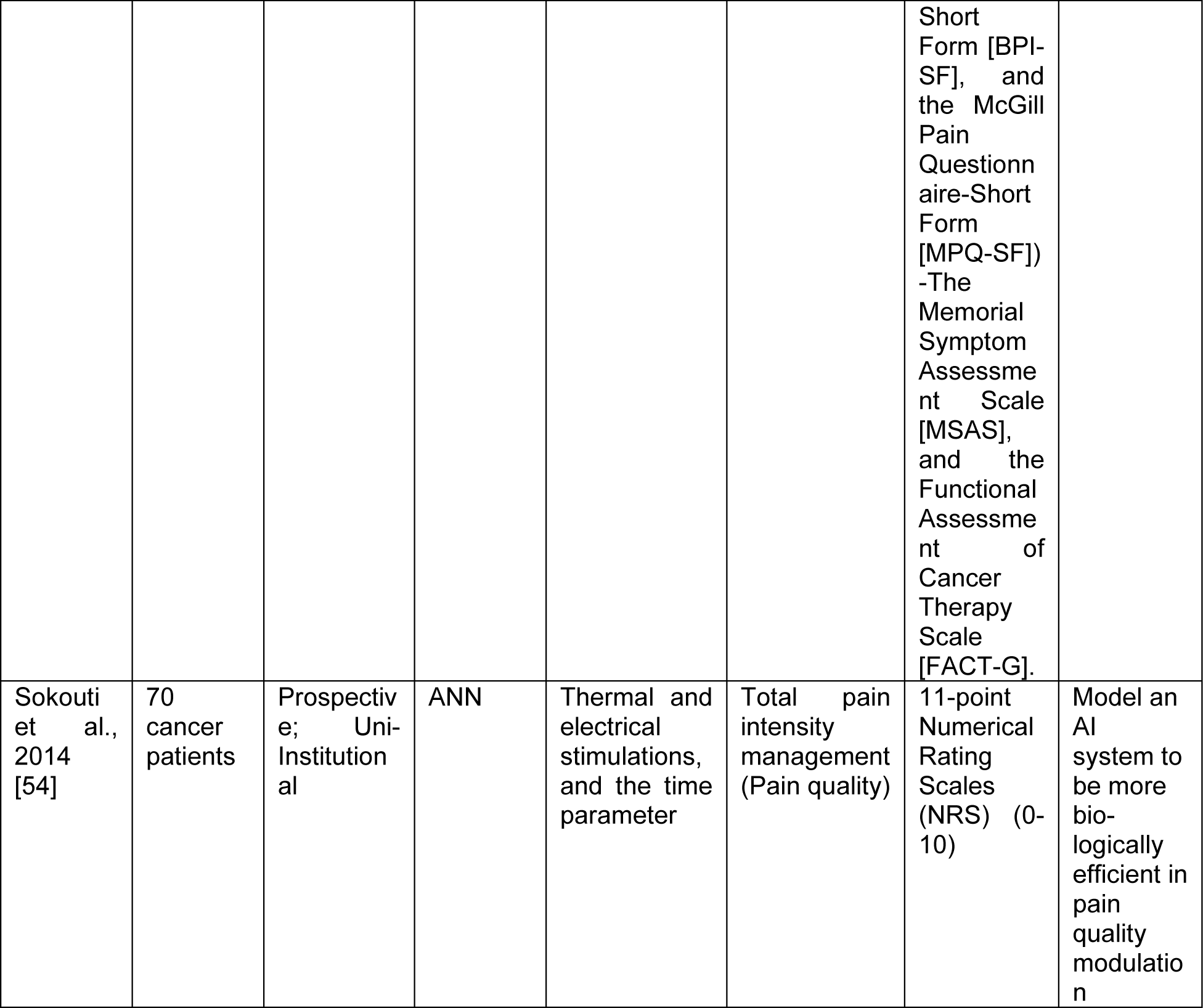
Characteristics of studies used AI/ML algorithms in cancer pain management prediction and decisions research.

Kumar et al. (2023) [34] to create and validate AI/ML predictive models for postoperative fentanyl analgesic demand and associated outcomes. Employing generalized linear regression models (GLM), Linear-SVM, RF, and Bayesian regularized neural network (BRNN), their research amalgamated nongenetic clinical and genetic factors (SNPs), cold pain test (CPT) scores, and pupillary response to fentanyl (PRF) to predict 24-hour postoperative fentanyl needs, pain scores, and time for the first analgesic. The models demonstrated varied R-squared values (0.313 for SVM—Linear, 0.434 for SVM—Linear, and 0.532 for RF), indicating moderately good predictive capacity for these outcomes. Olesen et al. (2018) [47] conducted a study to predict oral morphine equivalent dose (MED) in cancer patients using SVM based on 18 SNPs within genes linked to opioid receptors and metabolic pathways. The SVM analysis found no significant associations between the specific genetic variants and the required opioid dose in cancer pain patients. While Bobrova et al. (2020) [27] conducted a study involving 90 pancreatic cancer patients utilizing fentanyl transdermal therapeutic system (TTS) for pain relief. The research employed diverse ML models and examined 57 genetic and non-genetic factors to predict pharmaco-resistance of fentanyl. The study identified the SVM as the most effective model for classifying pharmaco-resistance. Facciorusso et al. (2019) [51] conducted a study with 156 pancreatic cancer patients undergoing repeat celiac plexus neurolysis (rCPN) using ANN and LR to predict pain response post-rCPN. The study found that the ANN outperformed the LR model in predicting treatment response. Dolendo et al. (2022) [45] analyzed 148 cancer patients who underwent mastectomy, utilizing LR to assess factors affecting postoperative opioid use. The study suggested that these models, if implemented, could significantly impact preoperative counseling and patient satisfaction by aiding in the prediction of postoperative pain. Im et al. (2011) [62] involved 428 ethnic minority cancer patients to evaluate a Decision Support Computer Program for Cancer Pain Management (DSCP-CA). The DSCP-CA was developed to assist nurses in managing cancer pain among ethnic minority patients, potentially enhancing care in this specific patient population. Sokouti et al. (2014) [54] created an ANN model for pain management using thermal and electrical stimulations and time parameters to address total pain intensity and pain quality. The main finding revealed the ANN’s accurate emulation of clinical data derived from stimulations, effectively managing severe pain in patients. A lack of external validation and clinical evaluation of the models was also detected in all studies involved in this subgroup.

### Performance of cancer related pain and pain management AI/ML models

The main discrimination performance evaluation metrics of the AI/ML models were accuracy, receiver operating curve-area under the curve (ROC-AUC), sensitivity, specificity and root mean square errors. Several studies did not report any outcome performance of the models used. Table 5 summarizes the discrimination performance results and includes an assessment on the validation methods used and whether model calibration was assessed for the studies that applied AI/ML models in cancer pain studies.

**Table 5:**
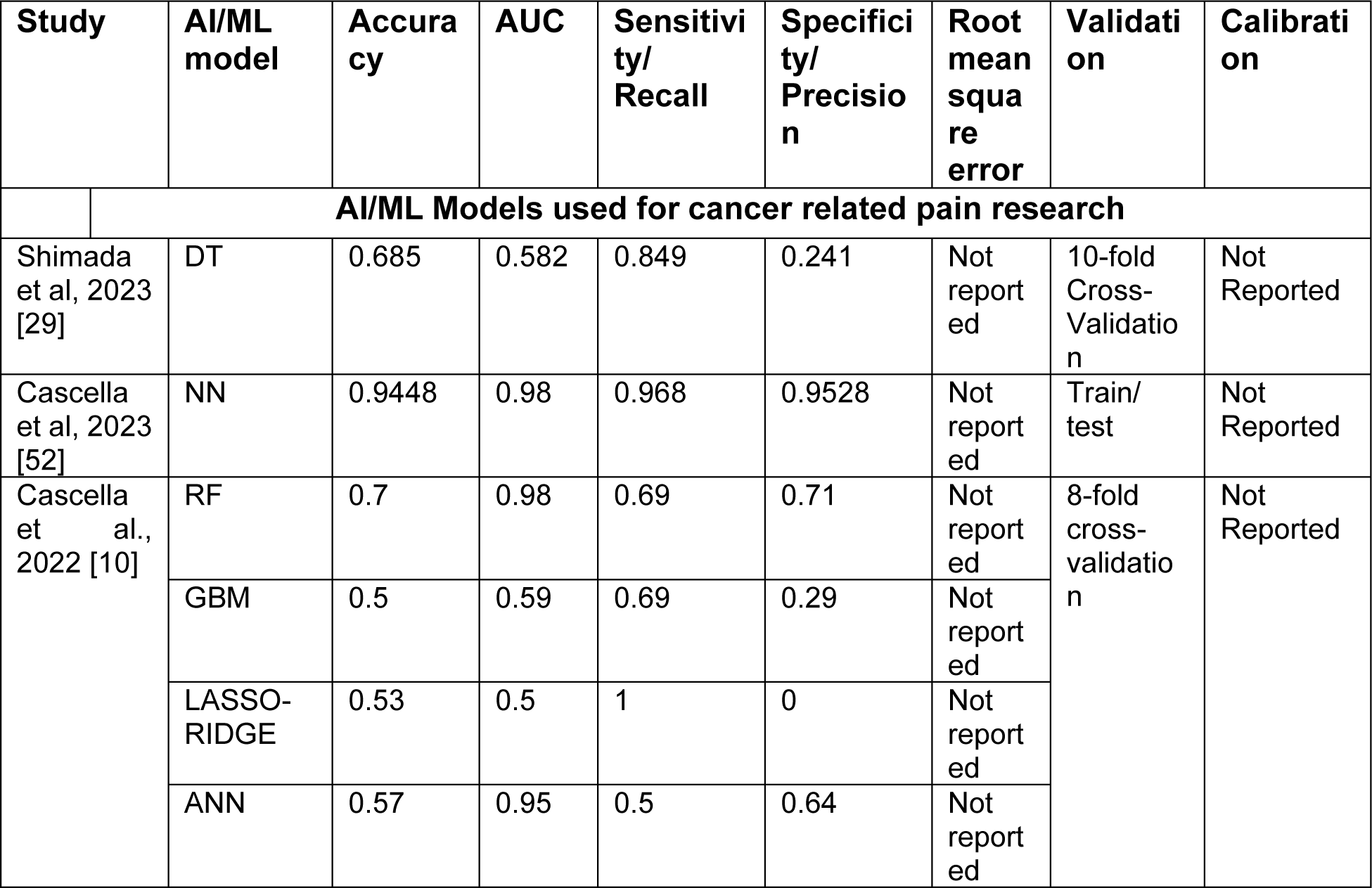
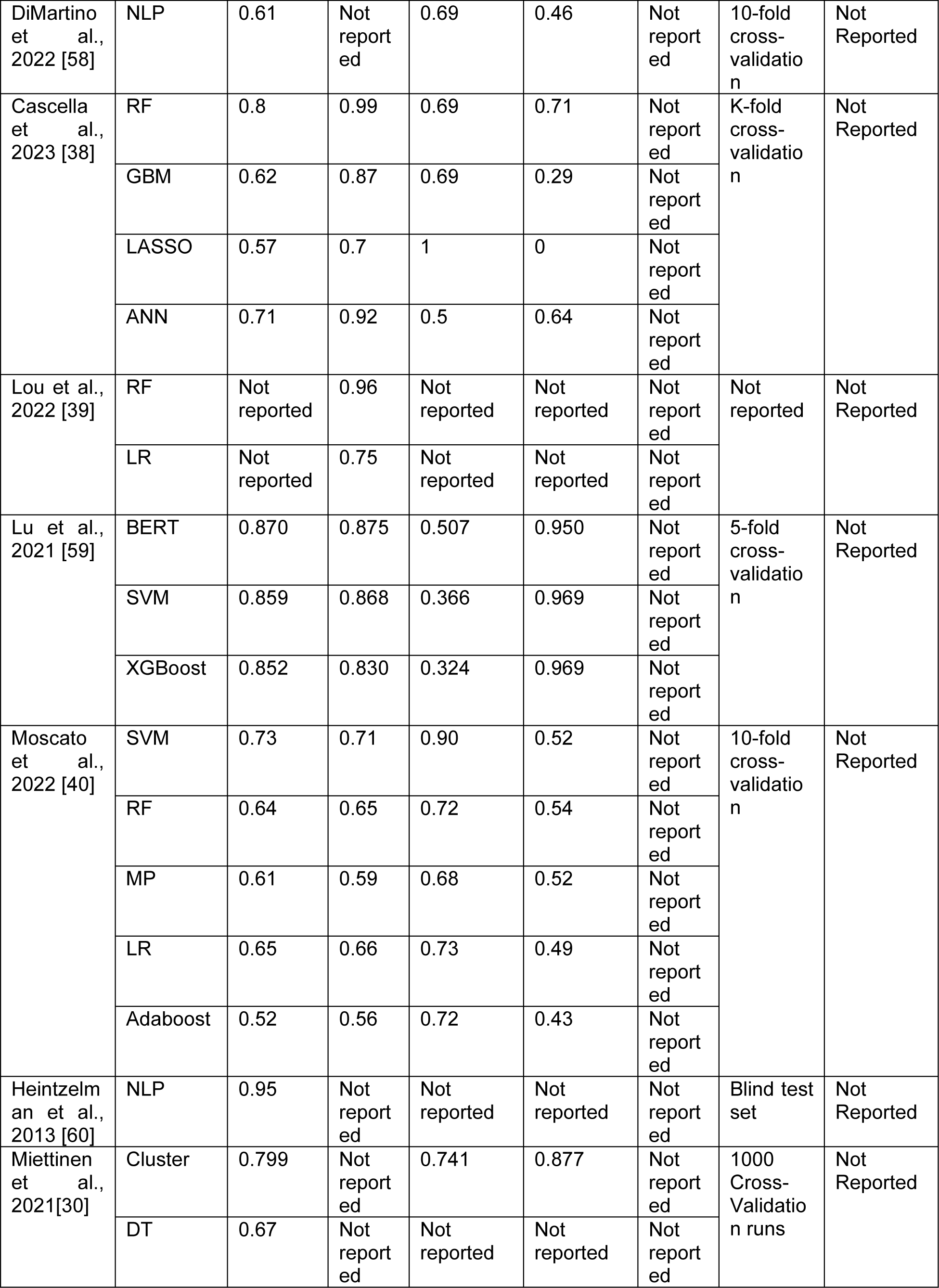
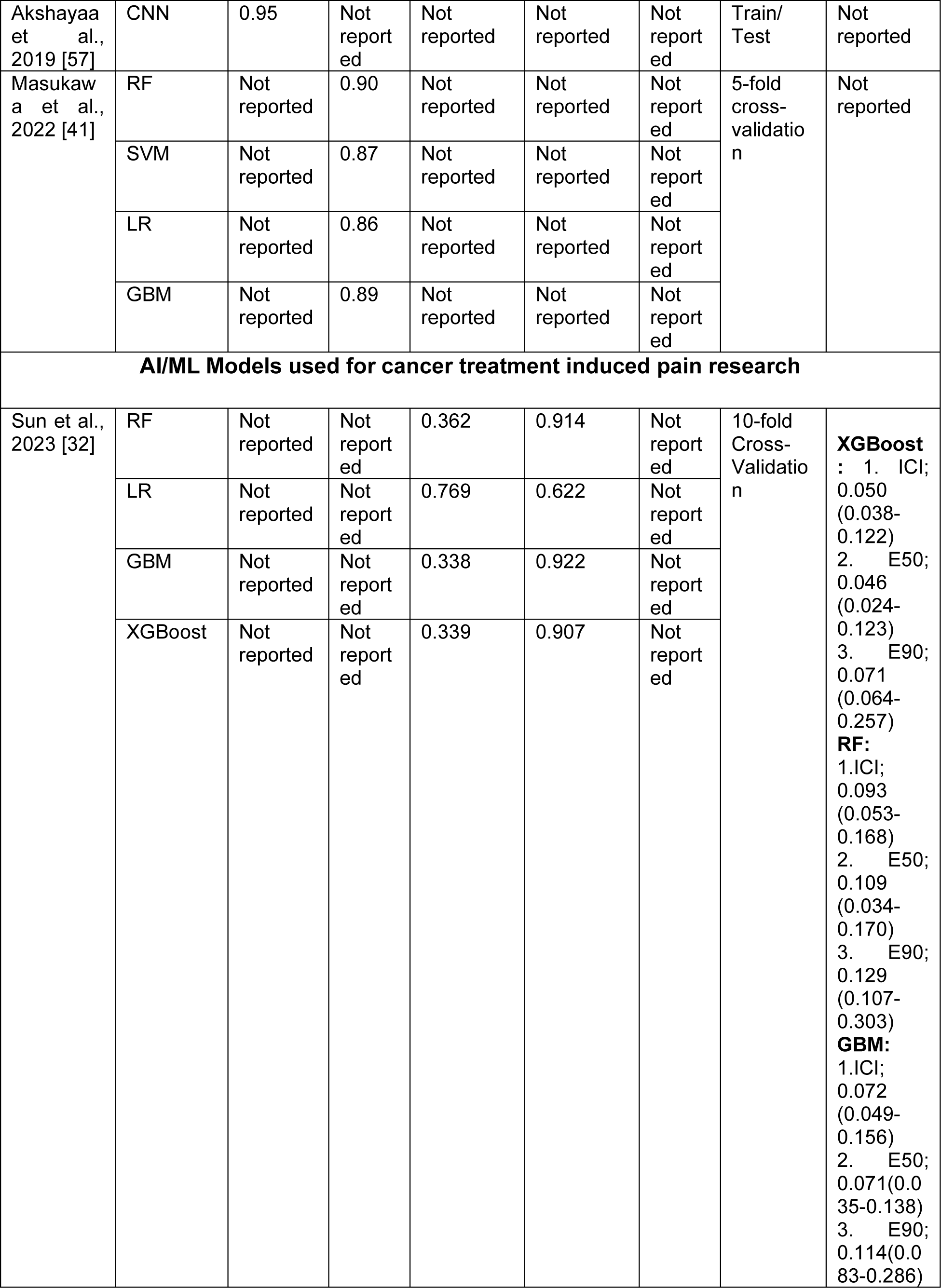
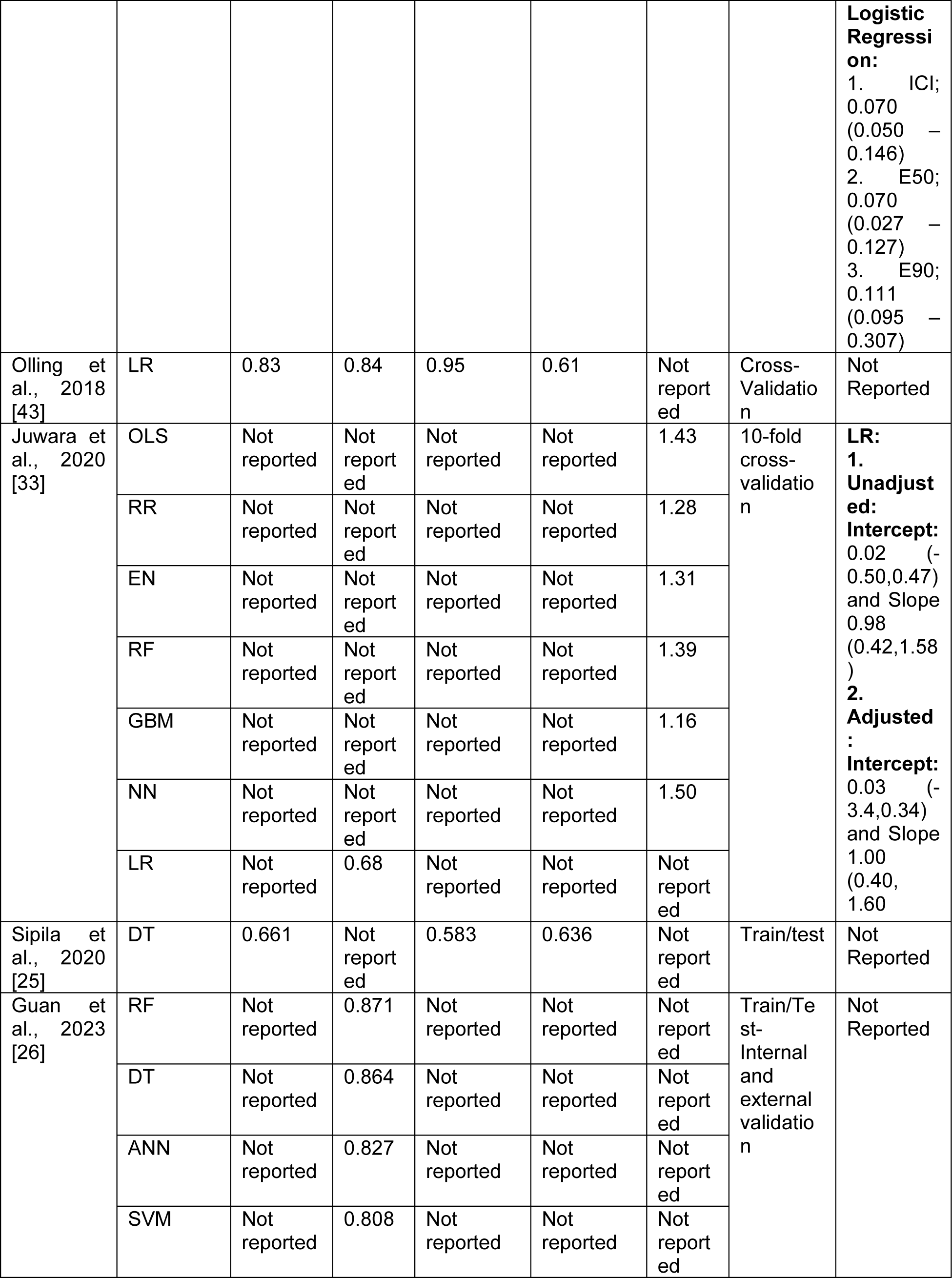
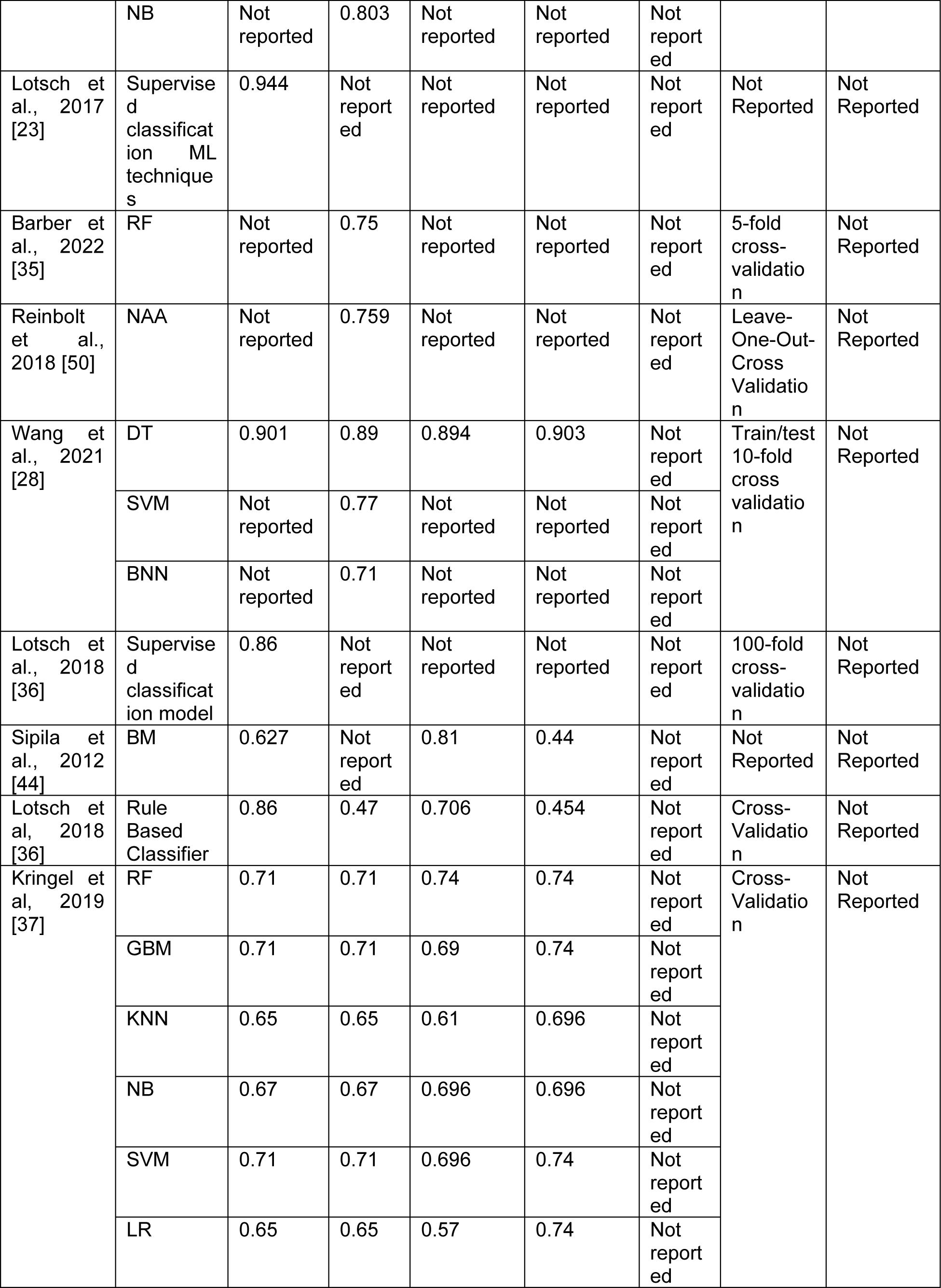
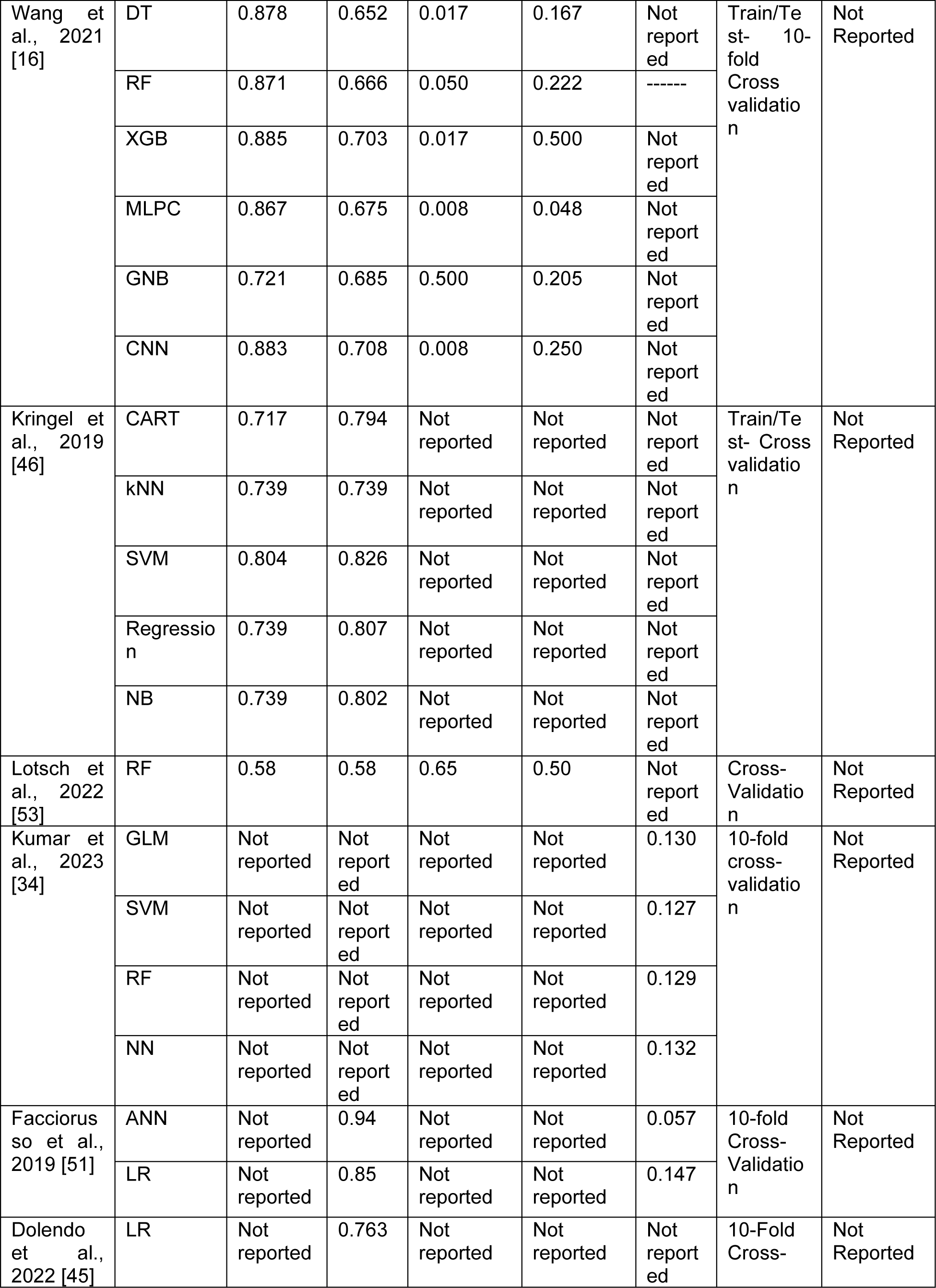
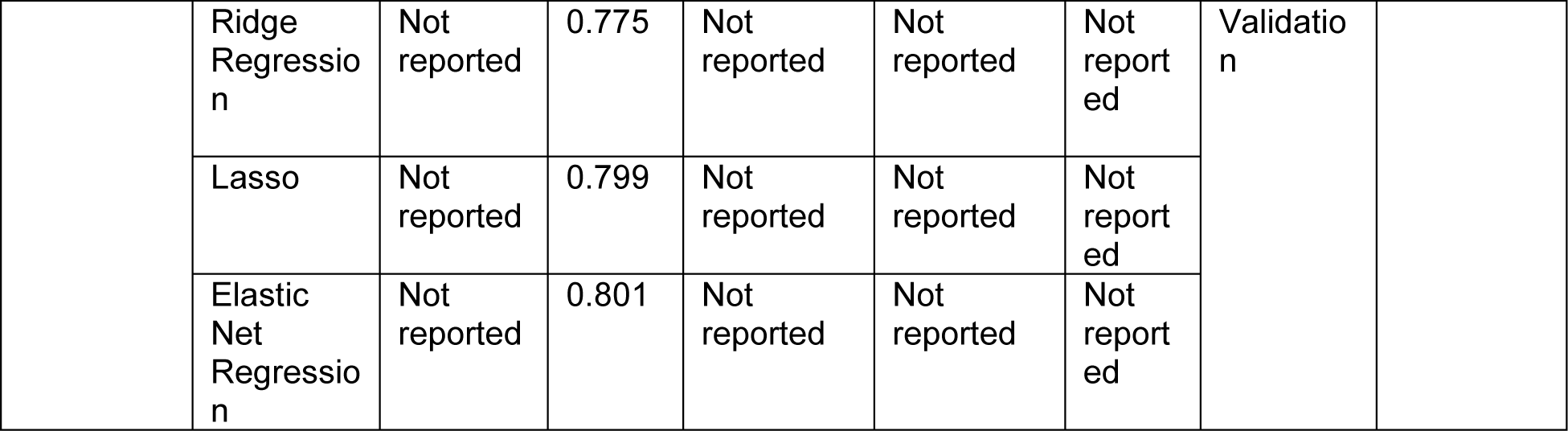
Models performance, validation and calibration reported in included studies.

Median AUC across studies reported the models AUC performance was 0.77% (range 0.47-0.99). Models used for cancer related pain studies showed the highest AUC with median AUC 0.86 (range 0.50-0.99), while the median AUC across studies of AI/ML models for cancer treatment related pain research and cancer pain management was 0.71 (range 0.47-0.89) and 0.80 (range 0.76-0.94), respectively (Figure 5.a). The RF model showed the highest median AUC (0.81, range 0.58-0.99), while the Lasso model showed the lowest median AUC (0.70, range 0.50-0.79). Median AUC for SVM (0.808, range 0.71-0.87), NB (0.80, range 0.67-0.803), LR (0.78, range 0.65-0.86), NN (0.79, range 0.65-0.98), DT (0.76, range 0.58-0.89) and boosting (GBM) (0.71, range 0.56-0.89) (Figure 5.b). The Lasso model demonstrated the highest sensitivity (1.00), while the lowest specificity (0.00). SVM demonstrated the highest median specificity (0.74, range 0.52-0.97).

**Figure 5:**
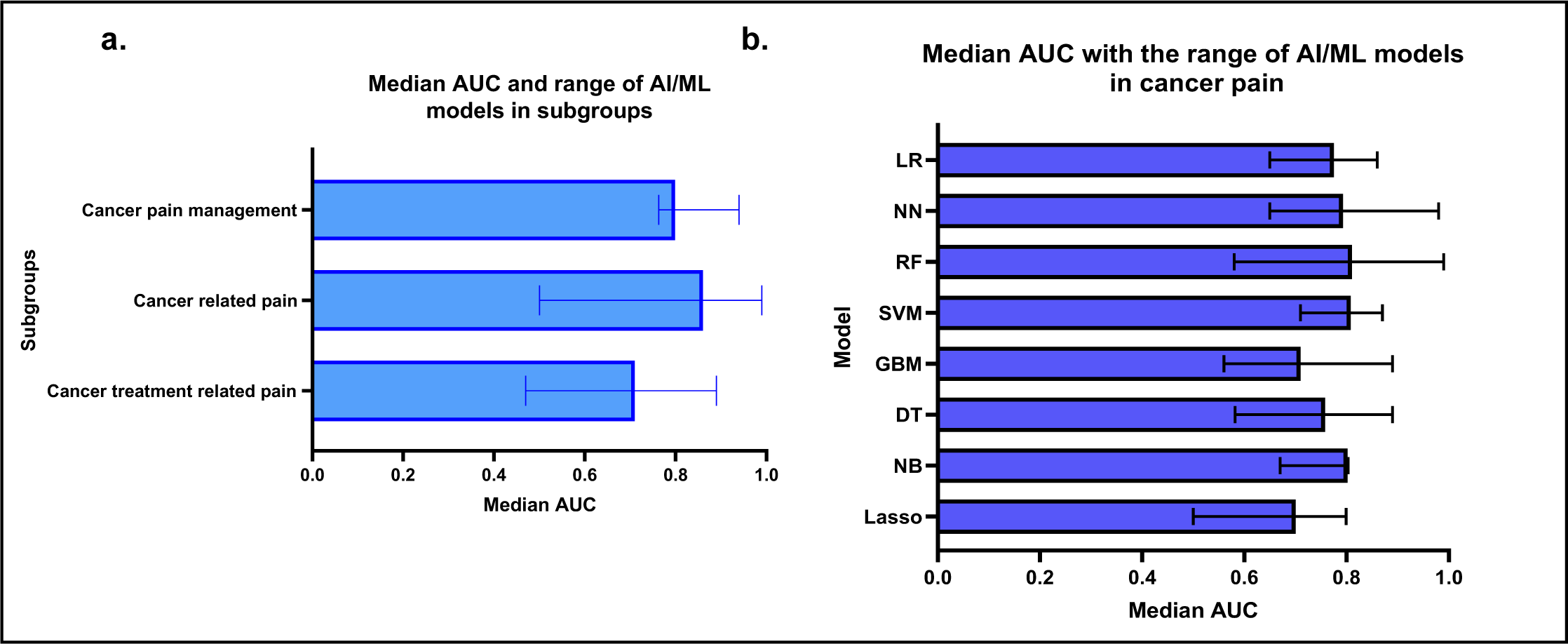
**a.** Median area under the receiver operating curve (AUC) across all included studies by subgroups. **b.** Median area under the receiver operating curve (AUC) across all included studies by AI/ML model.

Only 2 studies (Sun et al., 2023 [32] and Juwara et al., 2020 [33]), out of all articles reported the calibration of the AI/ML models (Table 5). Sun et al., used the integrated calibration index, E50, E90 and Hosmer-Lemeshow (H-L) test for calibration of the models used in the study, where XGBoost model showed better performance than multivariable LR model with ICI, 0.05 (95% CI (0.038-0.122)) which was lower than LR ICI, 0.07 (95% CI (0.050 – 0.146)) and the other models; RF and GBM didn’t show superiority in ICI and H-L P values reported for all models were (LR: 0.059, RF:0.429, GBM: 0.384 and XGBoost: 0.829) [32]. Jawara et al., generated the calibration plot of the predicted probability against the observed probabilities. The MCA adjusted model showed good calibration. The MCA unadjusted logistic classifier, the intercept: 0.02 (-0.50,0.47) and slope 0.98 (0.42,1.58), while for the MCA adjusted logistic classifier, the intercept: 0.03 (-3.4,0.34) and slope: 1.00 (0.40, 1.60) [33].

### Adherence to TRIPOD Guidelines

The overall compliance with the TRIPOD guidelines reporting checklist was 70.7%, revealing that 7 out of 31 domains fell below a 60% adherence rate. While reporting adherence surpassed 80% for components such as Study Design, Eligibility Criteria and Statistical Methods, it plummeted below 50% for crucial elements like blinding of outcomes/predictors, handling of missing data, model development, and identification of risk groups. Specifically, 75.4% of studies adequately specified their study population concerning inclusion/exclusion criteria and baseline characteristics. Lastly, 95.5% of abstracts provided adequate information on study methodology, and approximately 70% of studies disclosed funding sources (Figure 6).

**Figure 6:**
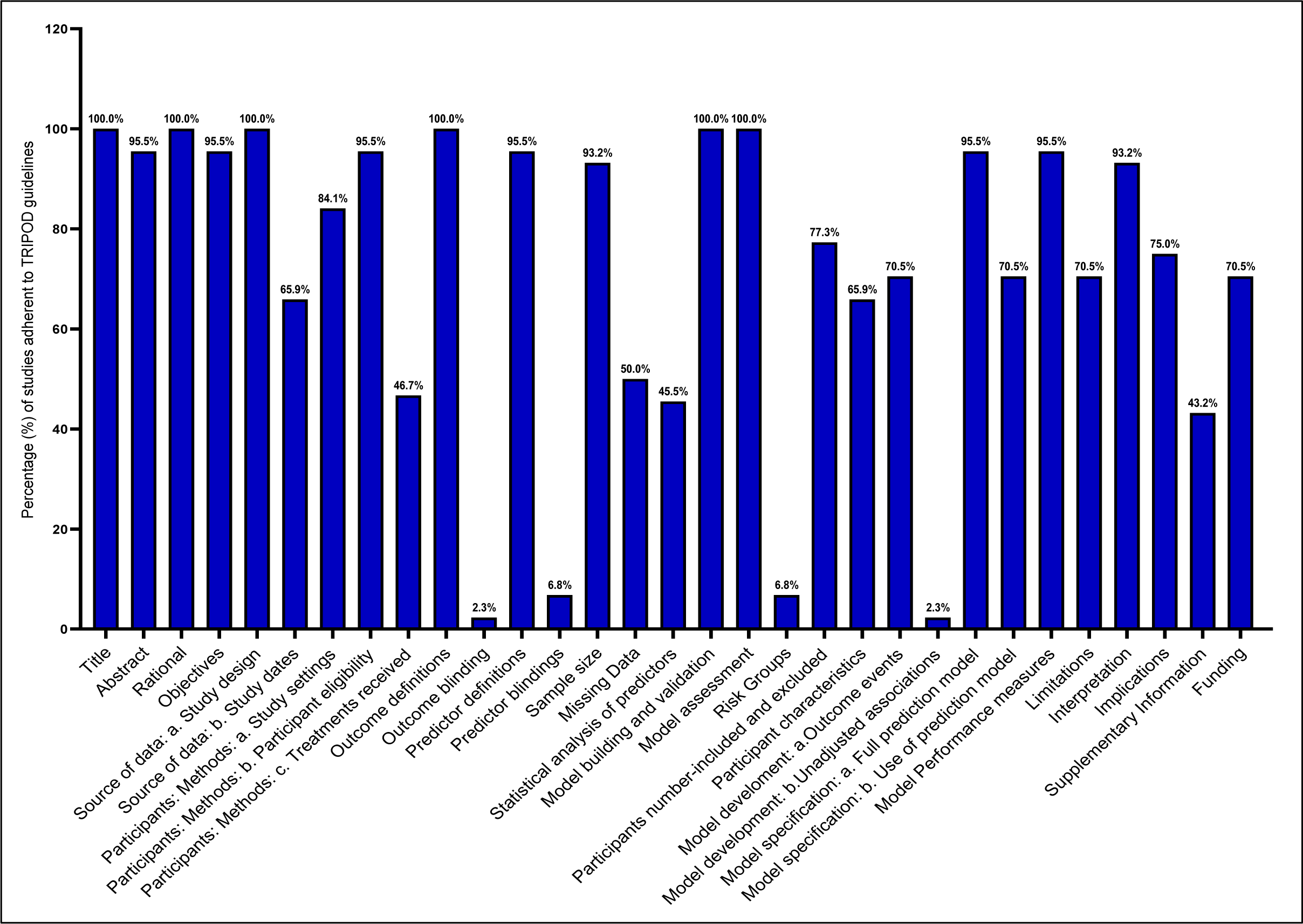
Frequency of adherence on included studies to reporting checklist of Transparent Reporting of a multivariable prediction model for Individual Prognosis or Diagnosis (TRIPOD) guidelines.

## Discussion

This comprehensive review delves into 44 studies that were identified through database searches, each utilizing distinct AI/ML methodologies within the field of cancer pain research published between 2006 and 2023. Most studies used prospective uni-institutial cohorts of cancer patients. In comparison to conventional statistical methods, AI/ML techniques have demonstrated superior performance, particularly in cancer pain prediction and pain management according to Sun et al. (2023) [32] and Juwara et al. (2020) [33]. The utilization of ML models, trained on big data sets of multiple features, has exhibited impressive mapping to select categories, classify patients and guide pain management decisions.

The results in this systematic review highlighted the substantial benefits of implementing AI/ML in cancer pain research, particularly in the context of predicting pain in cancer patients who have undergone cancer treatments. Out of the 44 studies, 19 focused on post-cancer treatment pain prediction, and 18 studies aimed at enhancing pain prediction and identifying high-risk factors associated with pain in cancer patients. Furthermore, our review pinpointed 7 studies that applied AI/ML for predicting cancer pain management and aiding in decisions regarding pain management (e.g., opioids usage).

Our comprehensive review illuminated the diverse array of AI/ML models deployed in the field of cancer pain research. Most of these models took the form of supervised classification or regression algorithms, including DT, RF, GBM, and LR. In addition, unsupervised clustering algorithms were applied to classify subgroups of pain patients and to identify key pain-related parameters. For making informed pain management decisions, advanced NNs were utilized and validated. AI-based decision support systems emerged as promising tools in guiding pain management choices. Most studies investigated multiple models (each model was tested separately) (55%), where RF and LR were the most common models used in these multiple models’ studies (n=15 studies for each). RF models demonstrated the highest performance across all studies (median AUC 81%), and Lasso models demonstrated the highest sensitivity (100%) while the lowest specificity (0%).

The expansion of clinical, genetic, and healthcare-related data has created an impetus for leveraging cutting-edge AI/ML techniques to harness this wealth of big data for the betterment of healthcare outcomes, especially for cancer patients contending with both the disease and its treatments. In our review, studies collectively demonstrated the potential of AI/ML models in predicting, assessing, and understanding pain among cancer patients, utilizing various inputs such as patient characteristics, imaging, video analysis, and clinical variables to improve pain management and remote consultations. While most studies used demographic and clinical data as input variables, only 4 studies included genetic data as inputs, Olesen et al., and Reinbolt et al., used single nucleotide polymorphisms (SNPs) data as inputs for the models [47, 50]. More novel approaches were observed with studies including image data (pathology, radiological, dosiomic data) as model inputs related to cancer pain (Akshayaa et al.[57], and Chao et al., [24]). Our systematic review did not provide specific details on the evaluation process of these features or how they were selected for model input. Deep learning offers the potential of analyzing high dimensional data (e.g., images) in combination with non-image data. This is an opportunity that future studies should take towards more comprehensive models in cancer pain prediction. Five studies used video recording, machine signals or tests results as input data of the models [23, 34, 40, 52, 54]. Text mining and NLP open the opportunities for future studies in cancer pain using texts and different non-structured data,

Numerous studies have explored the power of AI and ML to predict pain in various medical diseases. For instance, Matsangidou et al., (2021) conducted a comprehensive investigation into the utilization of AI and ML techniques for pain prediction, showcasing the potential of these advanced technologies to enhance the accuracy and effectiveness of pain prediction and prognosis [65]. Such research exemplifies the growing significance of AI and ML in improving patient care and healthcare outcomes, However, studies specifically addressing cancer-related pain are limited, and there is a lack of data regarding the calibration of models and adherence to TRIPOD guidelines. To our knowledge, this is the first study to do a comprehensive investigation and analysis of all up-to-date studies focused on applying AI/ML models in cancer pain medicine and in cancer pain management decisions. We analyzed the different models applied in these studies and the median model performance. We categorized the studies according to the use of the models into three subgroups: cancer pain research, cancer treatment related pain, and cancer treatment (e.g., opioids) decisions. Our data analysis demonstrated an increase in the trend of AI/ML studies in cancer pain in the last few years, and that AI/ML models showed high performance in cancer pain prediction, classification and management decisions.

In our review, we extracted and analyzed data regarding the adherence to The Transparent Reporting of a Multivariable Prediction Model for Individual Prognosis or Diagnosis (TRIPOD) guidelines, models calibration, external validation, and clinical application of the AI/ML models. Interestingly, our results revealed 70.7% overall compliance with the TRIPOD reporting checklist. However, few studies tested models’ calibration (5%), performed external validation of internally tested models (n=6, 14%) or discussed the clinical application of the validated models (23%). According to Van Calster et. al (2019) [66], the performance assessment of AI/ML models that estimate disease risks or predict health outcomes for clinical decision-making should involve evaluating model discrimination (e.g., calculating ROC-AUC) and model calibration as essential elements in the evaluation process [66]. While most studies on AI/ML models focus on discrimination and classification performance, calibration of models is often overlooked. Poor calibration of predictive algorithms can be misleading, leading to incorrect and potentially harmful clinical decisions [66]. Furthermore, adhering to the TRIPOD guidelines is essential in the context of AI/ML models, these guidelines provide a structured framework for transparent and comprehensive reporting, ensuring the reliability and reproducibility of predictive models [17, 18]. Equally crucial is the process of external validation, where models are rigorously tested in diverse and independent datasets to assess their generalizability and reliability beyond the initial training data. This step is vital in affirming the robustness of the models and their applicability to real-world scenarios [67]. Additionally, recurrent local validation should be considered in addition to external validation to test models’ reliability, safety and generalizability for clinical application, and Youssef et al., (2023) proposed the Machine Learning Operations-inspired paradigm for recurrent local validation of AI/ML models to maintain the validity of the models [68]. Furthermore, the clinical application of established ML models is of utmost importance to bridge the gap between research and practical healthcare settings. Understanding how these models perform in clinical practice enhances their utility and ensures informed decision-making. Several studies investigate robust AI/ML models in healthcare, however, few of these models have been clinically applied due to several limitations to translate AI/ML into clinical practices [69]. By prioritizing adherence to TRIPOD guidelines, conducting thorough repetitive local validations, external validations, and emphasizing the clinical application of ML models, the healthcare community can foster trust in AI-based tools, ultimately leading to improved patient outcomes and more effective healthcare interventions.

While deep reinforcement learning (RL) has exhibited remarkable efficacy in making morphine dosage decisions and optimizing pain management within the intensive care unit (ICU), as demonstrated by Lopez-Martinez et al. (2019) [70], our review, unfortunately, did not uncover any studies that had leveraged RL for the optimization of cancer pain management. This observation underscores a potential avenue for future research and development in the field, highlighting the need for exploring the application of RL techniques to enhance the management of pain in cancer patients.

Studies included in our review collectively demonstrate diverse applications of AI/ML models in cancer pain research, illuminating their potential in predicting, understanding patient satisfaction, identifying pain-related attributes, and managing pain in cancer patients across different age groups and healthcare settings. The identification of high-risk cancer pain patients and their associated risk factors is instrumental in stratifying patients according to their pain risk, which, in turn, informs the development of personalized pain management strategies. The integration of AI and ML in cancer pain prediction and patient risk stratification streamlines the analysis of complex big data with minimal human intervention, offering a promising path to improved outcomes and enhanced QOL for cancer patients suffering with pain. Furthermore, AI/ML techniques have proven effective in not only predicting but also diagnosing, categorizing, and recommending treatments for cancer pain. Although the clinical importance of applying these AI/ML models, several studies included in our review provided very limited details on the clinical validation of their AI/ML models in real healthcare settings, which is a crucial step in ensuring the applicability and reliability of these models in clinical practice. Clinical validation, in larger and more diverse cohorts, is vital to establish the predictive value of identified features and optimize pain treatment for cancer patients, enhancing the model’s real-world applicability, generalizability and effectiveness in clinical settings.

### Limitations

While our diligent effort involved an extensive comprehensive exploration of the implementation of AI and ML in the domain of cancer pain medicine, it is important to have some caution in interpreting the results due to the limitations in the design of our study. Notably, we encountered substantial heterogeneity among the identified studies, encompassing variations in the models employed, diverse AI/ML performance metrics, and disparities in the outputs associated with pain. Additionally, there is a lack of some AI/ML performance metrics which may affect the overall models’ performance. Furthermore, it’s essential to acknowledge that our search strategies did not explore other symptoms or cancer therapy toxicities that may have relevance to pain, such as mucositis, dysphagia, pneumonitis, and more. Our primary focus predominantly centered on patient-reported pain phenotypes, a deliberate choice that should be considered when evaluating the scope and implications of our findings. Very few data were available on the model calibration, external validation, and clinical application of the tested models in included articles which adds limitation of interpretation of the degree of biases, robustness, and clinical reliability of the applied models.

### Future Directions

Generalizability of the performance of the identified models that demonstrated high performance is required with external validation. Assessment of the robustness, clinical application of these models is needed to be conducted in the future for clinical use in real-world healthcare settings. The use of DL and RL models in the inclusion of imaging data for cancer pain medicine should be further explored.

### Conclusion

Recent advancements in AI and ML techniques have ushered in a new era of cancer pain research, with applications including cancer pain prediction, the anticipation of pain induced by cancer treatments, and aiding in pain management decisions. AI/ML models, showed high performance in predicting cancer pain, risk stratification, and enabling personalized pain management, with a median AUC 0.77 across all models and all studies. Although the adherence to TRIPOD guidelines was 70.7%, testing models’ calibration was 5%, the models’ external validation was 14% and the clinical application was 23%. There is an ongoing need for AL/ML models to be rigorously tested for calibration and externally clinically validated before their implementation in the real-world healthcare setting.

### Funding Statement

Drs. Moreno and Fuller received project related grant support from the National Institutes of Health (NIH)/ National Institute of Dental and Craniofacial Research (NIDCR) (Grants R21DE031082); Dr. Moreno received salary support from NIDCR (K01DE03052) and the National Cancer Institute (K12CA088084) during the project period. Dr. Fuller receives grant and infrastructure support from MD Anderson Cancer Center via: the Charles and Daneen Stiefel Center for Head and Neck Cancer Oropharyngeal Cancer Research Program; the Program in Image-guided Cancer Therapy; and the NIH/NCI Cancer Center Support Grant (CCSG) Radiation Oncology and Cancer Imaging Program (P30CA016672).Dr. Fuller has received unrelated direct industry grant/in-kind support, honoraria, and travel funding from Elekta AB. Vivian Salama was funded by Dr. Moreno’s fund supported from Paul Calabresi K12CA088084 Scholars Program. Kareem Wahid was supported by an Image Guided Cancer Therapy (IGCT) T32 Training Program Fellowship from T32CA261856”.

### Conflict of interest

Authors declare that they have no known competing commercial, financial interests or personal relationships that could be constructed as potential conflict of interest.

## Data Availability

All data produced in the present study are available upon reasonable request to the authors

## Acknowledgement

We thank The American Legion Auxiliary (ALA) for the ALA Fellowship in Cancer Research, 2022-2024, through The University of Texas, MD Anderson Cancer Center, UTHealth Houston Graduate School of Biomedical Sciences (GSBS). We thank The McWilliams School of Biomedical Informatics at UTHealth Houston, The Student Governance Organization (SGO) Student Excellence award.

## Notes

### Competing Interest Statement

The authors have declared no competing interest.

### Funding Statement

This study was funded by NIDCR (K01DE03052) and the National Cancer Institute (K12CA088084)

