## Supplemental Tables for "Artificial Intelligence and Machine Learning in Cancer Related Pain: A Systematic Review"

**Supplementary Table S1. Ovid MEDLINE search strategy**

| | |
|---|---|
| 1 | exp Neoplasms/ |
| 2 | exp Medical Oncology/ |
| 3 | (cancer* or carcinom* or tumor* or tumour* or neoplas* or malignan* or myeloma* or leuk?emia* or lymphoma* or sarcoma* or melanoma* or oncolog*).ti,ab,kf. |
| 4 | Oncology Nursing/ |
| 5 | or/1-4 [Cancer or oncology] |
| 6 | exp Pain/ |
| 7 | Pain Measurement/ |
| 8 | Pain Management/ |
| 9 | exp Analgesics, Opioid/ |
| 10 | (pain or analgesic* or opioid*).ti,kf. |
| 11 | or/6-10 |
| 12 | 5 and 11 |
| 13 | Cancer Pain/ |
| 14 | 12 or 13 |
| 15 | limit 14 to english language |
| 16 | (animals not (humans and animals)).sh. |
| 17 | 15 not 16 |
| 18 | (mice or mouse or murine or rat or rats or rodent or cells or "in vitro" or "cell line").ti. |
| 19 | 17 not 18 |
| 20 | exp Artificial Intelligence/ |
| 21 | ("machine learning" or "artificial intelligence" or "expert system*" or "deep learning" or "natural language processing" or "neural network*" or "fuzzy logic*").ti,ab,kf. |
| 22 | exp neural networks, computer/ |
| 23 | 20 or 21 or 22 |
| 24 | 19 and 23 |

**Supplementary Table S2. Ovid Embase search strategy**

| | |
|---|---|
| 1 | exp malignant neoplasm/ |
| 2 | (cancer* or carcinom* or tumor* or tumour* or neoplas* or malignan* or myeloma* or leuk?emia* or lymphoma* or sarcoma* or melanoma* or oncolog*).ti,ab. |
| 3 | 1 or 2 [cancer] |
| 4 | limit 3 to english language |
| 5 | Human/ |
| 6 | Nonhuman/ or ANIMAL/ or Animal Experiment/ |
| 7 | 6 not 5 |
| 8 | 4 not 7 |
| 9 | (mice or mouse or murine or rat or rats or rodent or cells or "in vitro" or "cell line").ti. |
| 10 | 8 not 9 [Cancer or oncology] |
| 11 | exp *pain/ |
| 12 | cancer pain/ |
| 13 | exp pain assessment/ |
| 14 | exp pain measurement/ |

| | |
|---|---|
| 15 | pain intensity/ |
| 16 | analgesia/ |
| 17 | (pain or analgesic* or opioid*).ti,kf. |
| 18 | or/11-17 |
| 19 | 10 and 18 |
| 20 | exp artificial intelligence/ |
| 21 | exp machine learning/ |
| 22 | expert system/ |
| 23 | natural language processing/ |
| 24 | ("machine learning" or "artificial intelligence" or "expert system*" or "deep learning" or "natural language processing" or "neural network*").ti,ab. |
| 25 | or/20-24 [machine learning] |
| 26 | 19 and 25 |
| 27 | conference abstract.pt. |
| 28 | 26 not 27 |
| 29 | (cancer* or carcinom* or tumor* or tumour* or neoplas* or malignan* or myeloma* or leuk?emia* or lymphoma* or sarcoma* or melanoma* or oncolog*).ti. |
| 30 | exp *malignant neoplasm/ |
| 31 | 29 or 30 |
| 32 | 28 and 31 |

**Supplementary Table S3. Clarivate Analytics Web of Science search strategy**

| | |
|---|---|
| 1 | TS=(pain NEAR/2 (measurement or management or intensity or score)) |
| 2 | TI=(pain or analgesic* or opioid*) OR AK=(pain or analgesic* or opioid*) OR KP=(pain or analgesic* or opioid*) |
| 3 | #1 OR #2 |
| 4 | TS=(cancer* or carcinom* or tumor* or tumour* or neoplas* or malignan* or myeloma* or leukaemia* or leukemia* or lymphoma* or sarcoma* or melanoma* or oncolog*) |
| 5 | TS=(("machine learning" or "artificial intelligence" or "expert system*" or "deep le rning" or "natural language processing" or "fuzzy logic*" or "neural network*" )) |
| 6 | #4 AND #5 |
| 7 | #3 AND #6 and Meeting Abstract (Exclude – Document Types) and English (Languages) |
| 8 | TI=(mice or mouse or murine or rat or rats or rodent or "in vitro" or "cell line") |
| 9 | 7 NOT #8 |
